# Ensemble forecasts of COVID-19 activity to support Australia’s pandemic response: 2020–22

**DOI:** 10.1101/2025.09.10.25335544

**Authors:** Robert Moss, Ruarai J. Tobin, Mitchell O’Hara-Wild, Adeshina I. Adekunle, Dennis Liu, Tobin South, Dylan J. Morris, Gerard E. Ryan, Tianxiao Hao, Aarathy Babu, Katharine L. Senior, James G. Wood, Nick Golding, Joshua V. Ross, Peter Dawson, Rob J. Hyndman, David J. Price, James M. McCaw, Freya M. Shearer

**Affiliations:** Melbourne School of Population and Global Health, The University of Melbourne; Department of Econometrics and Business Statistics, Monash University; Defence Science and Technology Group; School of Computer and Mathematical Sciences, The University of Adelaide; The Kids Research Institute Australia; School of Physics, Mathematics and Computing, University of Western Australia; School of Population Health, University of New South Wales; Department for Health and Wellbeing, Government of South Australia; South Australian Health and Medical Research Institute; Department of Infectious Diseases, The Peter Doherty Institute for Infection and Immunity; School of Mathematics and Statistics, The University of Melbourne

## Abstract

During the COVID-19 pandemic, many countries used real-time data analyses, predictive modelling, and COVID-19 case forecasts, to incorporate emerging evidence into their decisions. In Australia, national and jurisdictional public health responses were informed by weekly ensemble forecasts of daily COVID-19 case counts for each of Australia’s eight states and territories, produced by a consortium of researchers under contract with the Australian Government. We evaluated approximately 100,000 predictions for daily case counts 1–28 days into the future, generated between July 2020 and December 2022, and report here (a) how the ensemble forecasts supported public health responses; (b) how well the ensemble forecast performed, relative to the forecasts produced by each contributing team; and (c) how we refined our reporting and visualisations to ensure that outputs were interpreted appropriately. Similar to COVID-19 forecasting studies in other countries, we found that the ensemble forecast consistently out-performed the individual model forecasts, and that performance was lowest when there were rapid changes in the epidemiology, such as periods around epidemic peaks. Our consortium’s inter-nal peer-review process allowed us to explain how features of each ensemble forecast related to the design of the individual models, and this helped enable public health stakeholders to interpret the forecasts appropriately. Ultimately, our forecasts pro-vided information that supported public health responses during periods of different policy goals, and over a wide range of epidemic scenarios.

## 1 Introduction

The COVID-19 pandemic resulted in major social disruptions around the world [1, 2], as governments implemented a range of measures to curb infections and limit deaths in the face of a novel virus [3–5]. Initial decisions were guided by public health response plans, and as the pandemic unfolded governments increasingly made use of modelling to incorporate emerging evidence into their decisions [6–11]. Australia’s national and jurisdictional public health responses were informed by the results of real-time analyses that were presented to key government advisory committees in weekly situation reports, starting 4th April 2020 [12, 13]. Between July 2020 and December 2023, these reports included ensemble forecasts of daily COVID-19 case counts for each of Australia’s eight states and territories.

Beginning on 1st February 2020, the Australian government progressively closed borders to countries with established epidemics, culminating in almost complete cessation of travel from 20th March 2020, and imposed mandatory 14-day hotel quarantine on overseas arrivals from 16th March 2020. These measures were effective in limiting community exposure from infected international arrivals. Australia’s initial COVID-19 response focused on preventing local transmission [14] and, despite a steady influx of imported cases, local elimination was achieved for prolonged periods throughout 2020 and 2021, punctuated by several waves of local transmission. Australia’s COVID-19 vaccination program began in February 2021, with the aim of pivoting from preventing local transmission to “reopening” the country (i.e., easing non-pharmaceutical interventions). Vaccination targets and reopening plans were informed by modelling studies [15, 16]. Beginning on 1st November 2021, Australian jurisdictions progressively relaxed quarantine requirements for returning fully-vaccinated Australian residents and citizens, and subsequently for international travellers, with quarantine requirements fully removed as of 3rd March 2022. Over the 2022 calendar year Australia experienced substantial local transmission of multiple COVID-19 variants, with major waves of Omicron BA.1, BA.2, and BA.5, consistent with global epidemiology [17]. Many of these policy decisions were directly informed by our situation reports and the ensemble forecasts contained within.

Here we present an analysis of these ensemble forecasts, demonstrating how the ensem-ble consistently out-performed the individual models that were included in the ensemble, and how these forecasts supported public health responses over the 2020–2022 calendar years. These case forecasts also served as an input for COVID-19 hospital bed occupancy forecasts (December 2021 to December 2023) that further supported Australia’s public health responses [18]. A timeline of key events during this period is presented in Table 1.

**Table 1:** A timeline of key events prior to, and during, the study period (July 2020 to December 2022, inclusive)

| Year | Month | Event |
| --- | --- | --- |
| 2020 | Jan | First cases imported |
|  |  | National goal of preventing local transmission is announced |
|  | Mar | Limited local transmission occurs nationally |
|  |  | Hotel quarantine established for returning residents |
|  | Jun | Sustained local transmission occurs in Victoria |
|  | Oct | Local transmission is suppressed in Victoria |
| 2021 | Feb | Vaccine roll-out for priority and high-risk individuals begins |
|  | May | Vaccine roll-out for all adults begins |
|  | Jul | Delta variant becomes established in NSW (and subsequently in other jurisdictions) |
|  | Aug | National reopening plan targets are announced |
|  | Nov | NSW is the first jurisdiction to remove quarantine requirements for returning residents |
|  | Dec | Omicron BA.1 becomes established in all jurisdictions except WA |
| 2022 | Jan | Omicron BA.1 activity peaks in affected jurisdictions |
|  |  | Vaccine roll-out for children aged 5–11 begins |
|  |  | Booster doses recommended for at-risk individuals |
|  | Feb | Borders opened to all vaccinated travellers except in WA |
|  | Mar | Omicron BA.2 and BA.5 become established |
|  |  | WA is the last jurisdiction to remove quarantine requirements |
|  | Jul | Omicron BA.5 activity peaks |
|  | Sep | Multiple variants become established |
|  | Dec | Multiple variants activity peaks |

## 2 Methods

### 2.1 COVID-19 case data

For target data, we used de-identified line lists of reported cases for each Australian state and territory, extracted at each week of the study period from the National Notifiable Disease Surveillance System (NNDSS). The data included the jurisdiction (state or territory), the date of symptom onset (where available), the date when the case notification was received by the jurisdictional health department, and whether the infection was epidemiologically deemed to be acquired locally or overseas. We imputed missing symptom onset dates and estimated reporting delays using a time-varying delay distribution to characterise the duration between symptom onset and case notification [19]. Symptom onset dates were reported for 82% of cases in 2020. As Australia’s response pivoted to “reopening” the country, and daily case counts greatly increased, symptom onset reporting decreased to 48% of cases in 2021, and 38% in 2022. Nationally aggregated daily case counts are shown in Figure 1.

**Figure 1:**
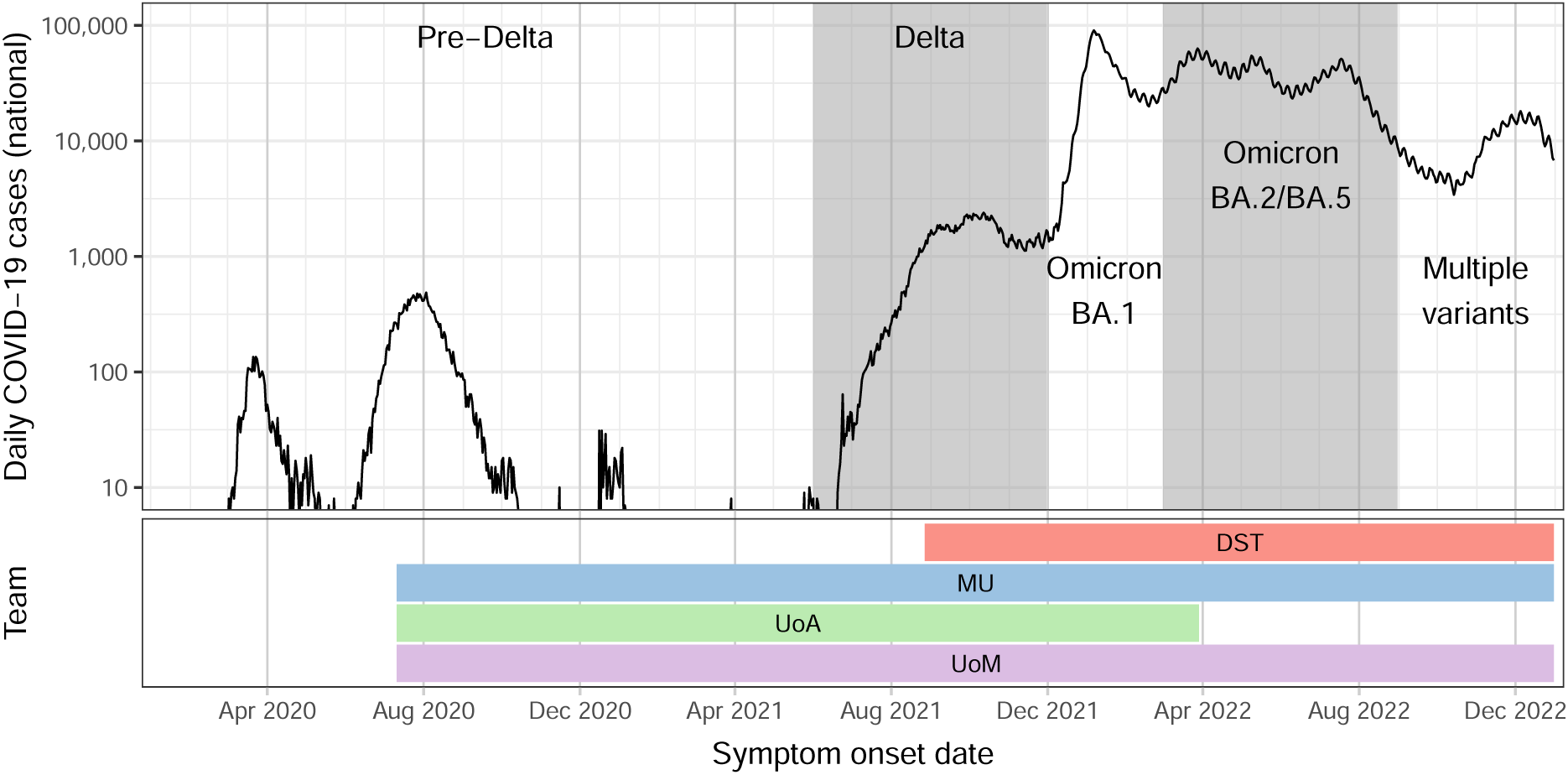
Top: Australian daily confirmed COVID-19 cases for the 2020–2022 calendar years. Dominant strain(s) are identified by alternating backgrounds (white and shaded), as determined by epidemiological assessment in situation reports [13]. Bottom: The periods over which each team contributed to the ensemble forecast. Team names and modelling approaches are listed in Table 2.

**Table 2:** The teams that contributed to the ensemble forecasts over the study period.

| Institute | Methodology |
| --- | --- |
| Defence Science Technology (DST) | Stochastic SEIR-type compartmental model |
| Monash University (MU) | Global autoregression model |
| University of Adelaide (UoA) | Branching process model |
| University of Melbourne (UoM) | Stochastic SEIR-type compartmental model |

### 2.2 Forecasting teams and models

Four modelling teams contributed forecasts for inclusion in the ensemble, using different types of model (see Table 2). This included two stochastic compartmental models, developed by the Defence Science and Technology Group (DST) and the University of Melbourne (UoM), which incorporated different assumptions about effective reproduction numbers and population immunity; a branching process model developed by the University of Adelaide (UoA); and an autoregressive model developed by Monash University (MU). We began with three teams contributing to the ensemble (MU, UoA, and UoM) and the DST team began participating in August 2021. After a four-month period where all four teams contributed to the ensemble forecast (2021-11-25 to 2022-03-22, inclusive), the UoA team stopped participating due to limited staff availability. Throughout this manuscript we will refer to the models, and the forecasts that they produced, using the acronyms listed in Table 2 and used in this paragraph.

The UoM team began contributing national forecasts in April 2020 [20], using a model adapted from a long-running seasonal influenza forecasting project [21–24], while the other teams created new models in response to the emergence of local COVID-19 transmission. All models were subject to ongoing development throughout the study, and these developments are documented in our weekly situation reports [13]. When new model features were introduced, retrospective forecasts were validated against previously-reported data before the new model iteration was considered for inclusion in the ensemble.

### 2.3 Inclusion/exclusion criteria

If there were known, or strongly suspected, data quality issues for any jurisdiction that could not be resolved with post-processing, we did not include ensemble forecasts for those jurisdictions in the weekly situation report. Models were also excluded when the forecasts produced for that week were assessed to be inappropriately sensitive to trends in the most recent case counts, which were known to be subject to time-varying reporting delays [19]. This decision-making process was facilitated by direct communication with data custodians in the relevant jurisdictions and national public health committees, which enabled rapid awareness of data quality issues.

Each week, the modelling teams generated forecasts from the provided data extracts and shared the results for peer review by the broader analysis team (comprising all modelling teams and other analysts, which we will refer to as the “consortium” hereafter) in a private online discussion board (in a Slack workspace). Peer review findings and retrospective performance results for each model were then tabled for discussion in weekly online consortium meetings, where we collectively reached agreement on which models, and model iterations, to include in the ensemble forecast for each jurisdiction that week (see Figure 2). This approach of ongoing discussion and reaching a collective consensus has been shown to improve the performance of human predictions in domains such as geopolitics [25].

**Figure 2:**
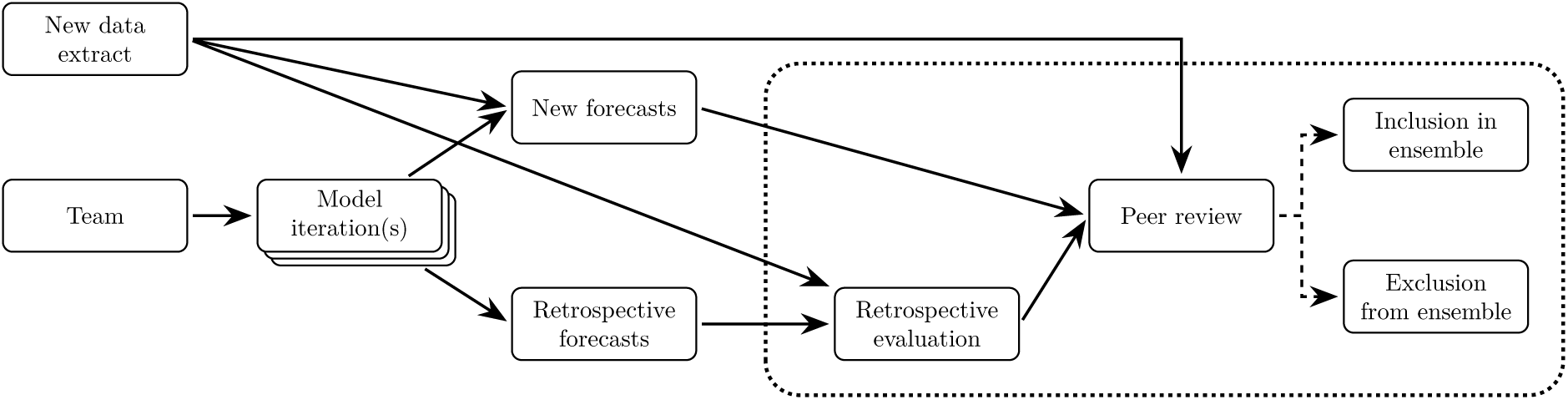
A flowchart of the weekly peer review and inclusion/exclusion decision process, shown here for the model iteration(s) provided by a single modelling team in a given week. The dotted box indicates activities and decisions made by the entire consortium.

Models were excluded when their outputs differed dramatically from the consortium’s collective expectations and the current epidemiological assessment, or when a technical issue was identified by the modelling team or by the peer review process. Importantly, each model was evaluated against the most recent data rather than competing against the other models (e.g., via benchmarks such as skill scores), with the purpose of deciding whether the model outputs were likely to be a reliable basis for decision-making [26]. The outcomes of these collective decisions are captured in publicly available reports [13].

### 2.4 Ensemble forecast and evaluation

Model forecasts were submitted in the form of 2000 simulated trajectories of daily case counts over the 28-day forecast horizon for each of the eight jurisdictions. We generated equal-weight ensemble forecasts for each jurisdiction by sampling trajectories from each model forecast for that jurisdiction. Notably, weights were allocated to each team rather than to each model iteration. For example, when a team contributed forecasts from multiple model iterations, we considered the iterations as representing a single model over a discrete parameter space. Accordingly, the addition or removal of one model iteration impacted the relative weights of all model iterations from that team, but did not affect the weights allocated to other teams.

As described in the previous section, we held weekly meetings in which we reviewed individual model forecasts and the ensemble forecast prior to including these forecasts in our weekly situation reports to government. Forecast performance was evaluated visually (using daily credible intervals) against previous weeks’ forecasts and the most recent data, and quantitatively using Continuous Ranked Probability Scores (CRPS). The weekly situation reports included plots of the ensemble forecast for each jurisdiction, and comparisons of the previous forecasts (made 1–4 weeks ago) against the most recent data for each jurisdiction. These comparisons included visualisations for the ensemble, and also for each individual model. In this study we also report forecast bias *B*(*t*) as per Funk et al. [27]:

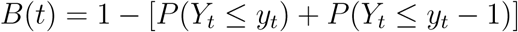

### 2.5 Data availability

The analysis code, summary outputs, figures, and supplementary materials are provided in a public git repository: https://gitlab.unimelb.edu.au/rgmoss/analysis-australian-covid19-ensemble-forecasts-results.

Note that this repository *does not include* the input cases files and output ensemble forecasts (approximately 2.3 Gb). These are provided in a separate data repository: doi:10.26188/29434298.

## 3 Results

We begin with an overview of the model inclusion/exclusion outcomes for each ensemble forecast. We then present an overview of the ensemble forecast performance, and demonstrate that the ensemble forecasts were more reliable than the forecasts generated by each individual model. We conclude this section by detailing how the ensemble forecasts supported public health activities during each phase of the study period (defined by predominant circulation of different COVID-19 variants, as illustrated in Figure 1) and highlight ways in which we improved forecast visualisations and reporting over the study period.

### 3.1 Models included in each ensemble forecast

As shown in Table 3, all models were regularly included in the ensemble forecasts. The DST and UoM teams often contributed forecasts generated from multiple model iterations, and when models were excluded from the ensemble it was most often *a single model iteration* from either team. The most common reason for exclusion was for a model to exhibit large and systematic biases in the forecasts produced in previous weeks (i.e., retrospective posterior predictive checks). And in several instances the UoA team encountered computational issues and were unable to provide forecasts in a timely manner.

**Table 3:** The percentage of ensemble forecasts for which each model was included, and the number of models included in each ensemble forecast (minimum, maximum, and mean), reported separately for each period in which particular strain(s) dominated. Note that models could be excluded from the ensemble for various reasons, from being assessed as inappropriately sensitive to trends in the most recent data, or otherwise producing suspect predictions, to being unavailable due to technical issues and/or delays in model development (see subsection 2.3).

| Strain | Forecasts | DST | MU | UoA | UoM | # Models |
| --- | --- | --- | --- | --- | --- | --- |
| Pre-Delta | 302 | — | 100.0% | 93.7% | 100.0% | 2–3 (2.99) |
| Delta | 183 | 4.4% | 100.0% | 100.0% | 100.0% | 3–4 (3.04) |
| Omicron BA.1 | 94 | 84.0% | 100.0% | 57.4% | 100.0% | 2–4 (3.41) |
| Omicron BA.2/BA.5 | 208 | 96.2% | 96.2% | 7.7% | 100.0% | 2–4 (3.00) |
| Multiple variants | 89 | 100.0% | 100.0% | — | 98.9% | 2–3 (2.99) |
|  | 876 | 376 | 868 | 536 | 875 | 2–4 (3.03) |

### 3.2 Summary of forecast performance

Ensemble forecasts for the months of March, April, and May 2022 are shown in Figure 3 for four of the eight Australian jurisdictions (forecasts for the remaining jurisdictions are shown in figures S3 and S4 in appendix S1). These example forecasts demonstrate key features that were exhibited by the ensemble forecasts over the entire study period (2020– 2022). In brief, the ensemble was generally in good agreement with the data. Consistent with findings from COVID-19 forecasting studies in other countries [6–11, 28, 29], it was an ongoing challenge to predict the timing of change points. The forecasts exhibited a tendency to overshoot peaks, and to undershoot the onset and ending of individual waves. In particular, the forecasts tended to predict substantially larger peaks than were ultimately observed. However, as shown in the 4-week forecasts in Figure 3, the observed peaks still fell within the forecast credible intervals. The extremely broad intervals for the Victorian forecast for 15 March 2022 arise from a single model, which was assessed as highly uncertain but consistent with the data, and included in the ensemble that week.

**Figure 3:**
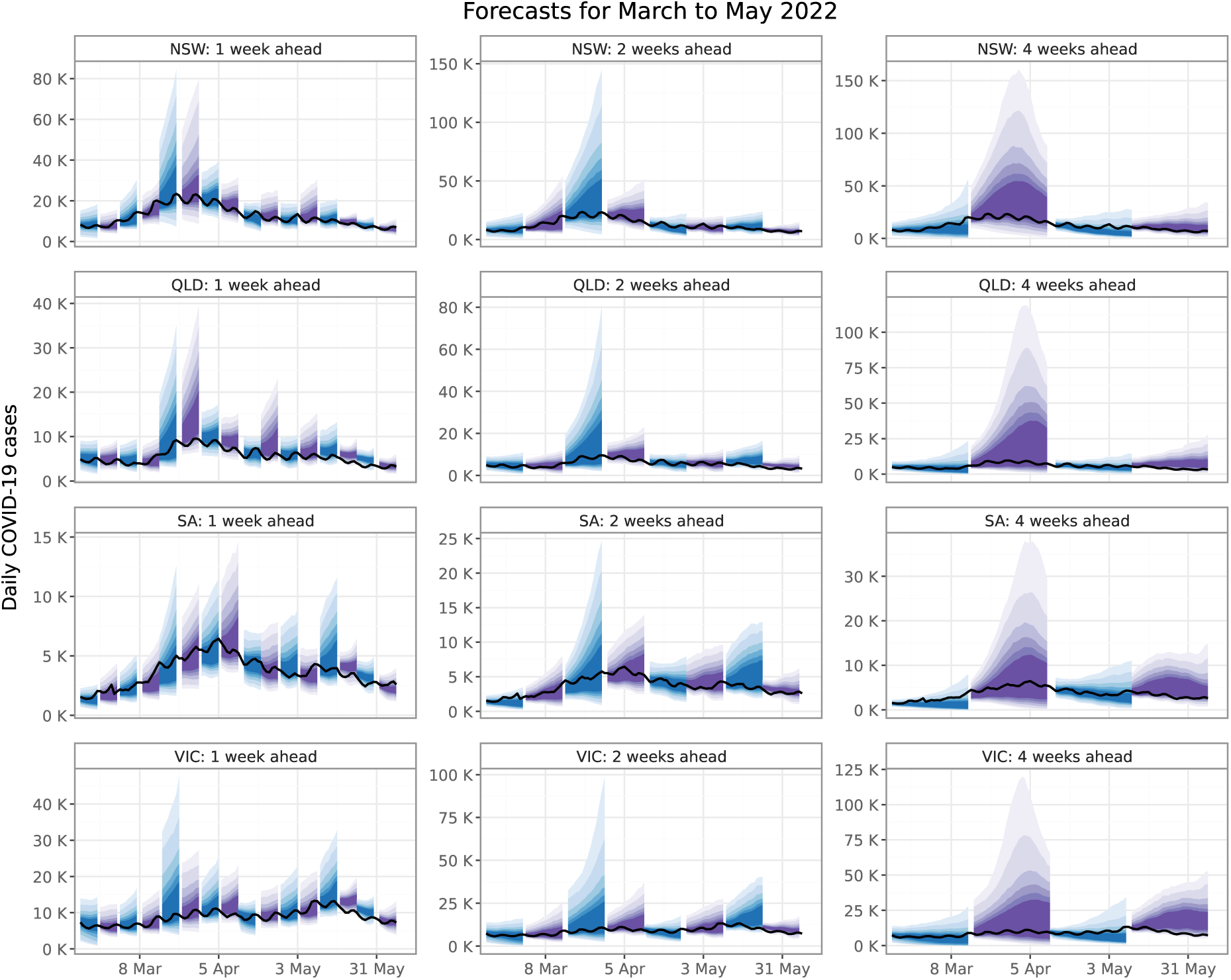
Ensemble forecasts over the months of March, April, and May 2022, shown for four Australian jurisdictions: New South Wales (NSW); Queensland (QLD); South Australia (SA); and Victoria (VIC). The left columns shows 1-week forecasts for each ensemble forecast, the middle column shows 2-week forecasts for every second ensemble forecast, and the right column shows 4-week forecasts for every fourth ensemble forecast. Shaded regions illustrate the 50%, 60%, 70%, 80%, 90%, and 95% credible intervals, and black lines depict the data as reported at the end of the study period.

Ensemble forecast coverage (shown in Figure 4) was reasonable across most pandemic waves. Coverage is the probability that a credible interval will contain the true value, and ideally an X% credible interval will include the true value X% of the time. The best coverage was observed during the “Pre-Delta” and “Delta” periods where, for example, almost exactly 50% of the ensemble 50% credible intervals included the true value. During this time, Australian jurisdictions responded rapidly to local disease activity in pursuit of strong suppression [14], and while the precise impact of these interventions was challenging to predict, there was only limited local COVID-19 transmission and case counts remained low. Australia then pivoted from pursuing strong suppression to reopening and accepting substantial levels of local COVID-19 transmission. In the subsequent “Omicron BA.1” period, the models failed to anticipate the massive surge and rapid decline in daily case counts; forecasts were overly confident and exhibited a tendency to first undershoot and then overshoot the true case counts. Forecast coverage improved markedly for the “Omicron BA.2/BA.5” and “Multiple variants” periods, with a slight tendency towards being overly uncertain. For periods where the individual models exhibited marked dif-ferences in forecast coverage, the ensemble forecast coverage was similar to, if not better than, the coverage of the best individual model.

**Figure 4:**
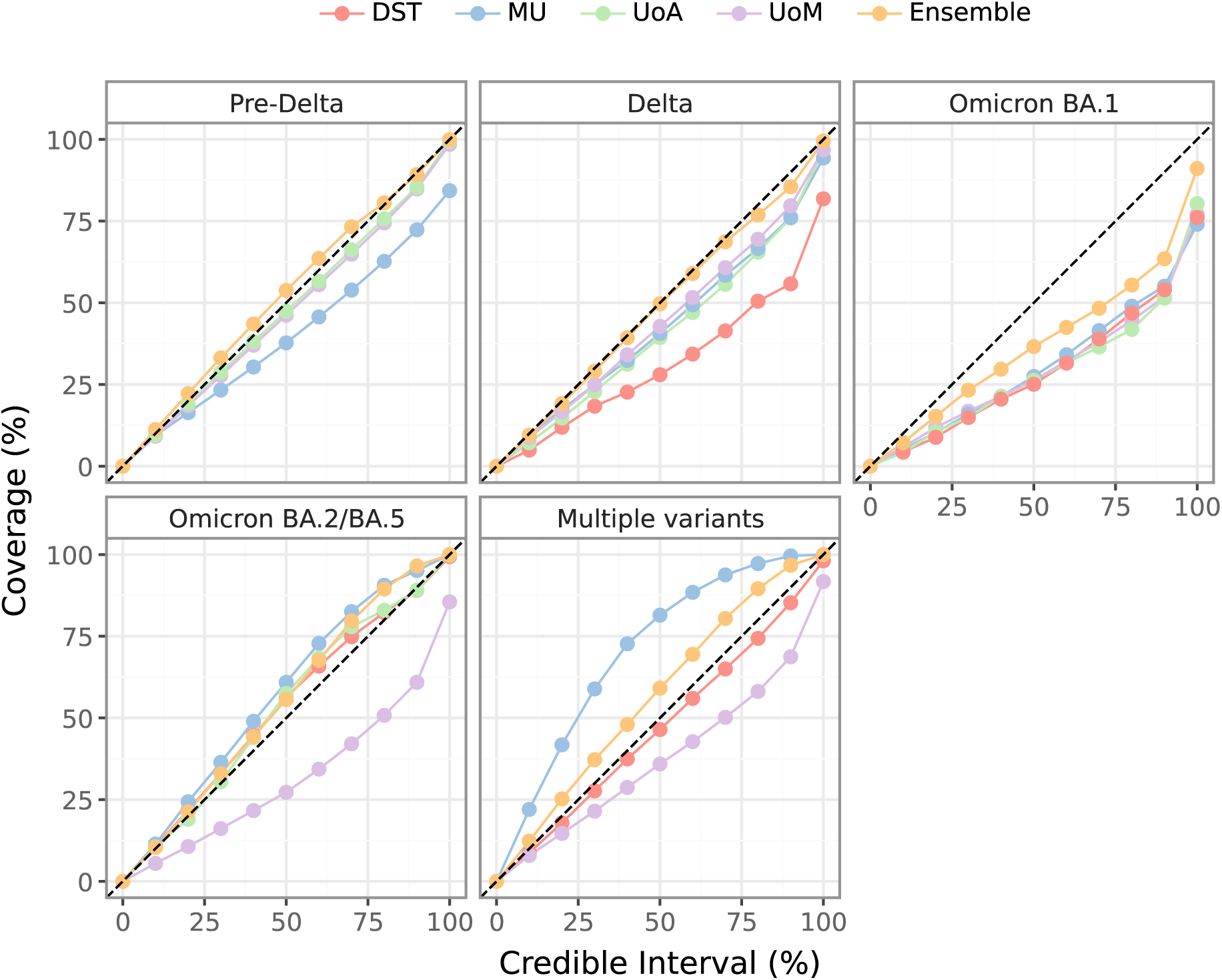
Observed coverage of the forecast credible intervals for each individual model, and for the ensemble, with respect to the ground truth case counts as reported after the end of the study period. Results are reported with respect to each dominating strain (refer to Figure 1). Dashed lines indicate perfect coverage. Points below the dashed lines indicate overly certain forecasts, points above the dashed lines indicate overly uncertain forecasts.

As might be expected, observations that lay further into the future were harder to predict. Forecast performance was highest for short lead times, and steadily decreased as the lead time increased. This was true not only on aggregate, but also for the majority of individual observations. As shown in Table 4, for a given observation the forecast performance consistently improved from the initial prediction (when the observation was 22–28 days ahead) to the final prediction (when the observation was 1–7 days ahead).

**Table 4:**
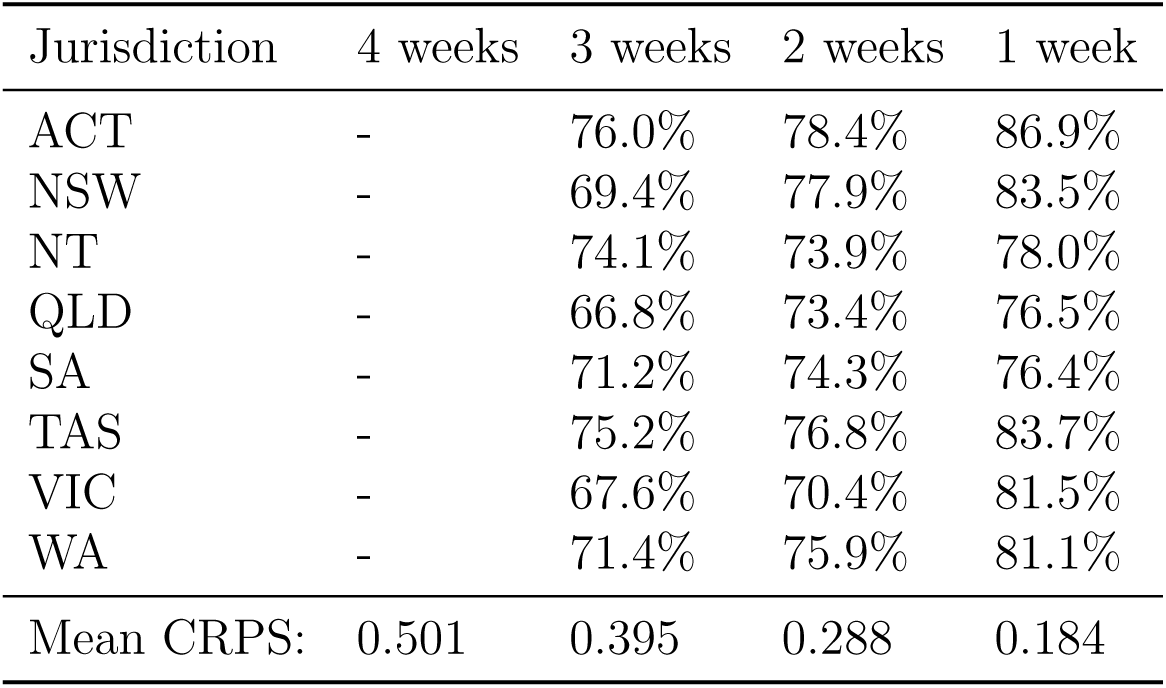
The improvement in ensemble forecast CRPS for a given observation, as the lead time becomes shorter (i.e., as the forecasting date approaches the observation date). Each cell shows the percentage of predicted distributions for which the CRPS decreased, relative to the predictions made one week earlier. The bottom row shows the mean ensemble forecast CRPS by lead time (calculated for all forecasts, not only those where CRPS decreased).

Forecasts exhibited substantial biases around epidemic peaks. This is illustrated in Figure 5, which shows the bias for each model and the ensemble for forecasts generated in the 4 weeks leading up to, and the 4 weeks after, each observed peak. Forecasts generated 2–4 weeks prior to observed peaks undershot the data (negative bias) more often than they overshot the data (positive bias). In contrast, forecasts generated from 1 week prior to 1 week after the observed peaks had a tendency to overshoot the data (positive bias), which highlights the challenge of identifying peaks in real-time when surveillance data are subject to ascertainment biases and reporting delays [12].

**Figure 5:**
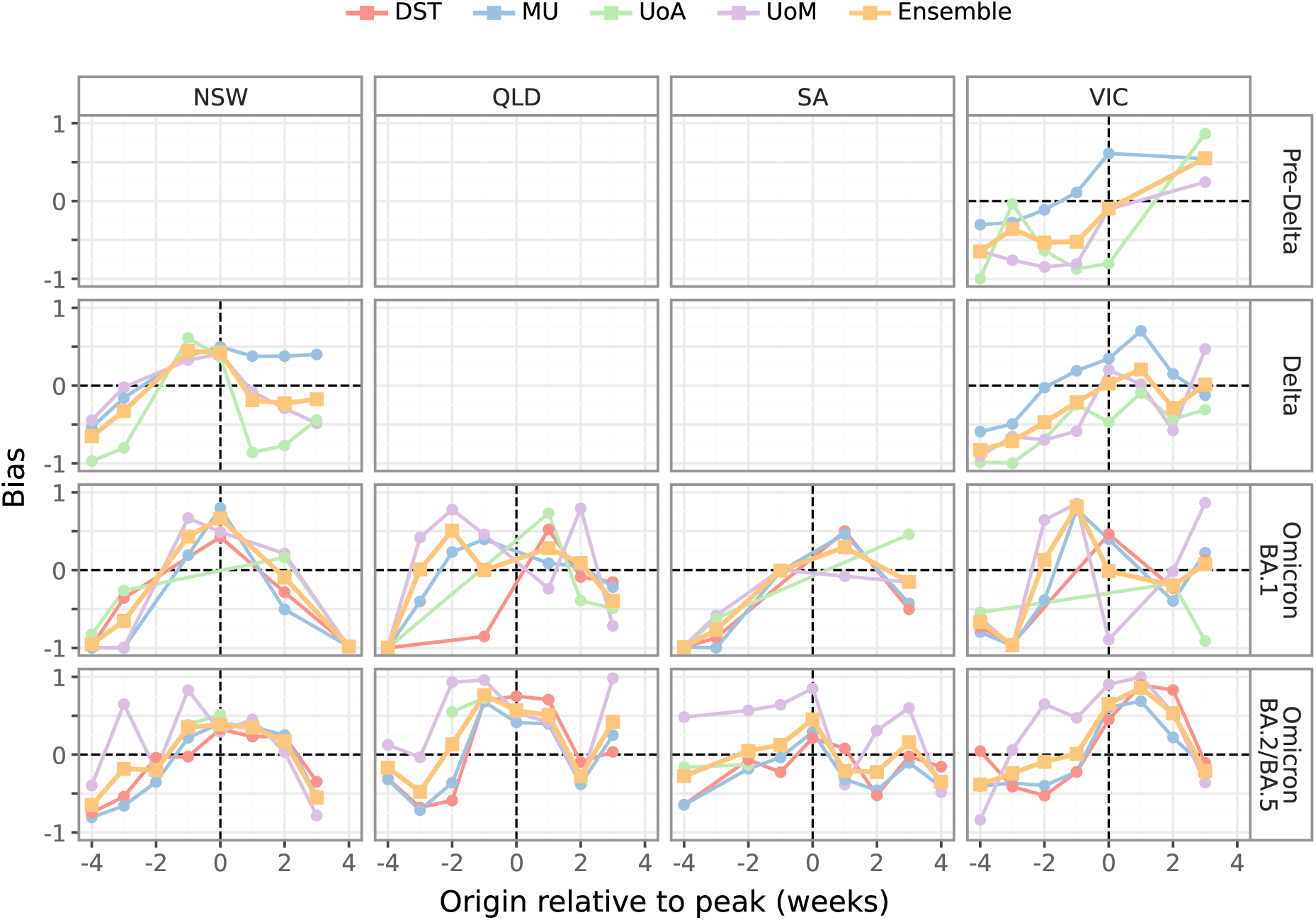
Forecast bias for each model and the ensemble, shown for forecast origins within 4 weeks either side of the largest peak (minimum 200 cases) observed in four jurisdictions (columns) for each dominant strain (rows). Positive bias indicates a tendency to overshoot the data, negative bias indicates a tendency to undershoot the data. The “Multiple variants” period is not included in the figure, because case counts were generally flat and the observed peaks occurred towards the end of the study period. See appendix S5 for forecast bias around these peaks over all eight jurisdictions.

No model consistently produced the least biased forecasts for weeks preceding or following the observed peaks. When there was a wide spread in bias, the ensemble forecast was often less biased than most, if not all, of the individual model forecasts. Similar trends are evident in individual model and ensemble forecast performance (see appendix S5); no model consistently produced the most accurate forecasts for weeks preceding or following the observed peaks, and the ensemble tended to mostly out-perform individual models.

While the ensemble forecasts struggled to accurately predict peak sizes and timing (evident in the measure of forecast bias, discussed above), in most weeks preceding or following an observed peak, a proportion of the forecast trajectories were positively correlated with the pre-peak and post-peak case counts (appendix S5). This indicates that the forecasts could at least partially characterise the qualitative trends in future case counts (i.e., whether case counts would increase, decrease, or remain stable).

### 3.3 Relative model performance

Recall that at each week of the study period, sample trajectories of daily case counts were provided over four-week horizons for each Australian jurisdiction. For each individual daily case count prediction, we ranked the individual models and the ensemble using CRPS on log-transformed values [30]. The results for days where at least one case was reported are shown in Figure 6. Collectively, the individual models were more likely than the ensemble to be the top-ranked forecast, but no single model dominated the top ranking. The strength of the ensemble was that it was *most often the best or second-best* forecast. Skill scores for each model, relative to the ensemble, further reinforce the finding that no model consistently out-performed the ensemble (Figure 7). The model rankings for each ensemble forecast (appendix S2) also demonstrate that there were no obvious predictors of which model would perform the best for a given time period or epidemiological context.

**Figure 6:**
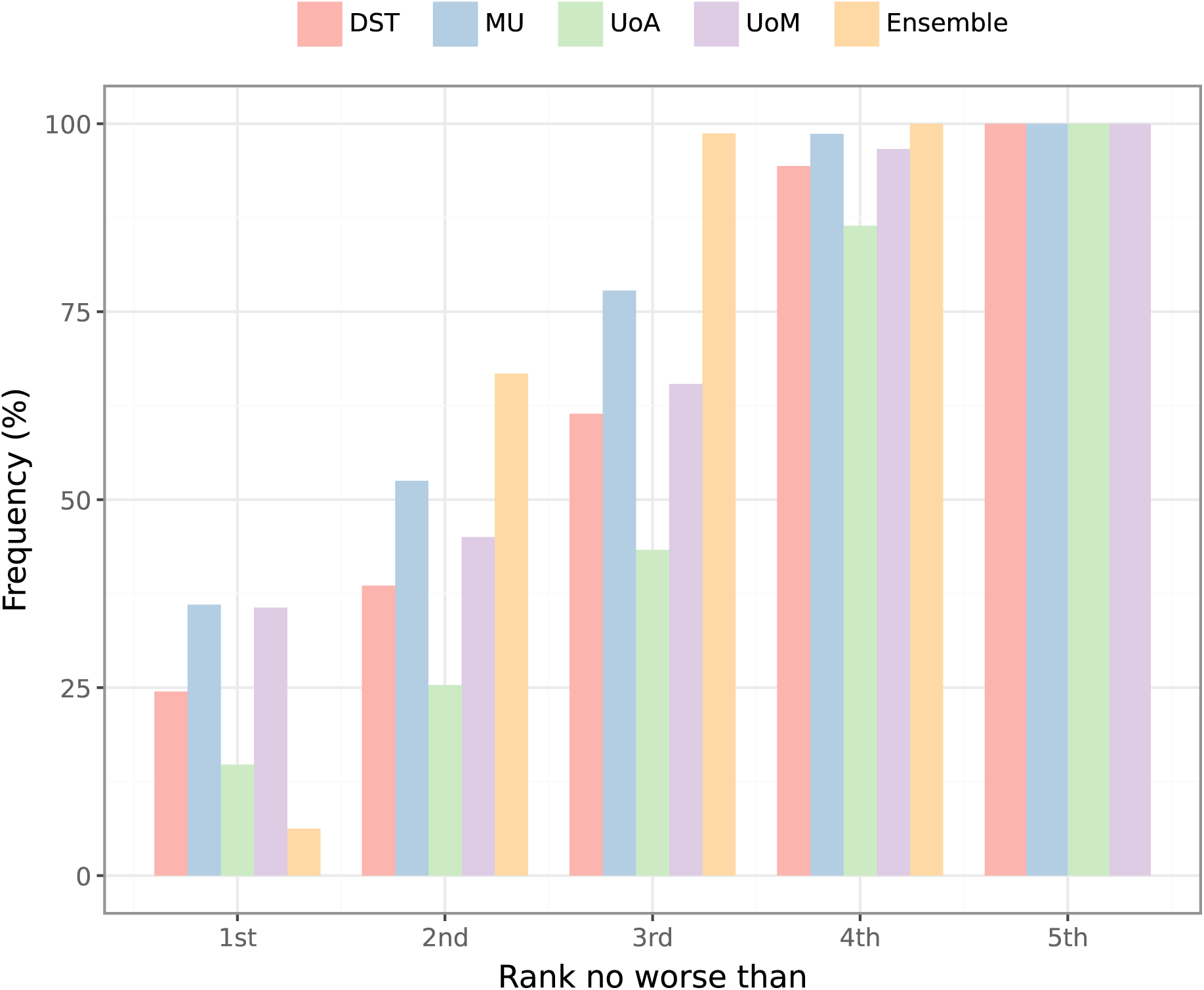
Cumulative rankings for each individual model and for the ensemble, for days where at least one case was reported, calculated using CRPS on log-transformed values. While the ensemble was rarely the top-ranked model, it was *at least* 2nd-best approximately two-thirds of the time (66.7%), and was *almost always* in the top three (98.7%).

**Figure 7:**
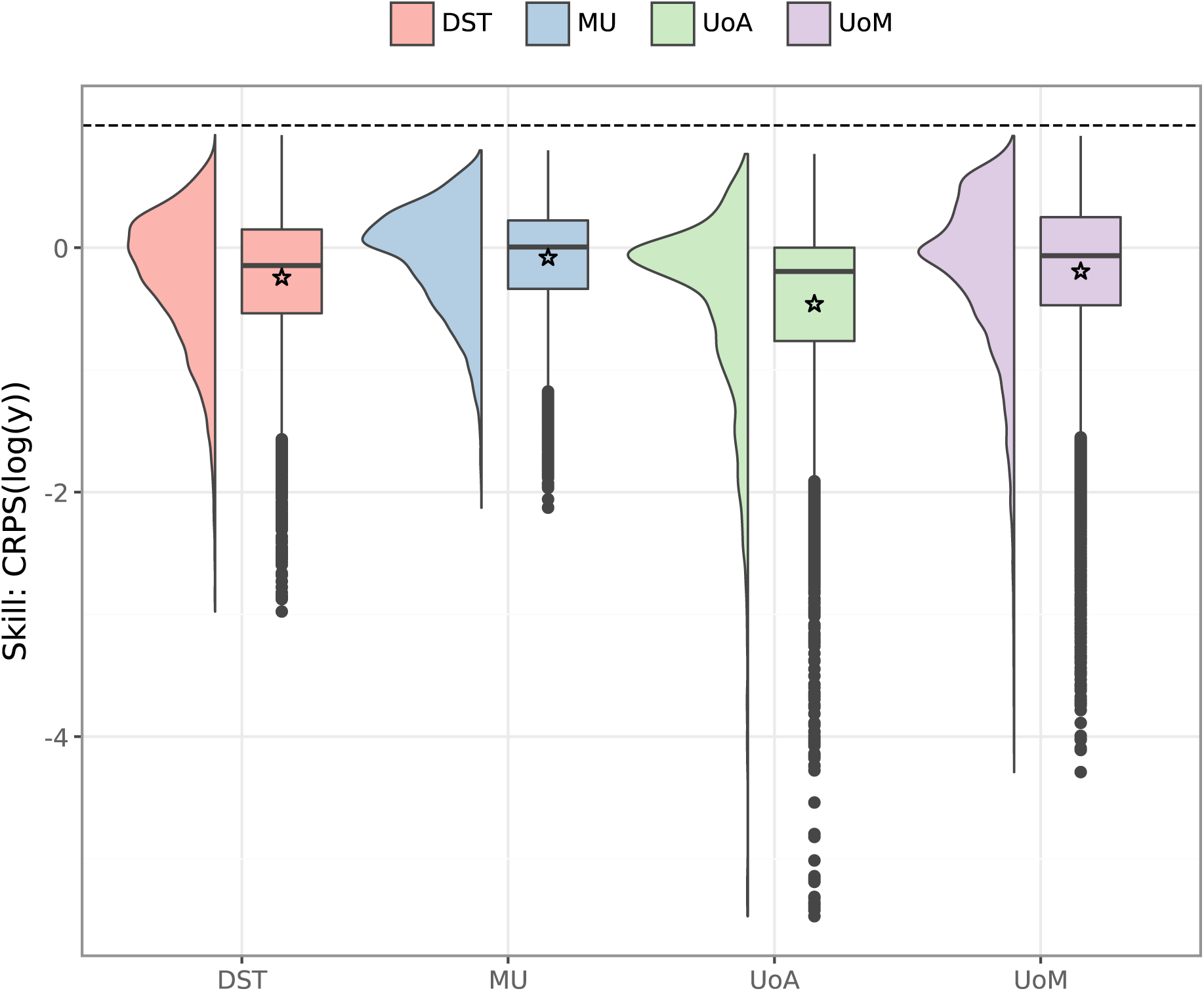
Distribution of skill scores for each individual model, measured against the ensemble, for days where at least one case was reported (around 53% of all observations). Skill scores were calculated using CRPS on log-transformed values [30]. Stars indicate mean skill scores. The MU model performed better than the other models, with a median skill score of 0.002. Mechanistic models performed well for extended periods of zero cases (median skill scores of 0.75–0.97, not shown here) but were otherwise outperformed by the ensemble, with median scores of *−*0.07 (UoM), *−*0.14 (DST), and *−*0.19 (UoA).

### 3.4 Pre-Delta: May to October 2020

We now highlight how the forecasts supported public health activities in each phase of the study period, beginning with the Pre-Delta phase. This first period of sustained local COVID-19 transmission began in mid-2020, where the majority of cases occurred in the state of Victoria. Ensemble forecasts for this period are presented in appendix S1. The performance of the UoM forecasts over this second wave has already been reported [12], and we begin by comparing the ensemble forecasts to the UoM forecasts over this wave. In the weeks prior to the peak in daily cases (3 August 2020), the upper bounds of the UoM forecasts substantially overshot the true peak, and predicted sustained exponential growth in cases. This exponential growth was tempered in the ensemble, which averaged over three models (MU, UoA, and UoM). Accordingly, the ensemble outperformed the UoM forecasts (which had a mean skill score of −0.37, see appendix S4), even though the ensemble forecasts tended to undershoot the data. By late July, we identified that case ascertainment and reporting delays had substantially increased, and our right-truncation adjustments were under-estimating the true case counts in the most recent days prior to each forecast. In the weeks after the peak, the UoM forecasts confidently predicted a sustained decrease in cases, while the MU and UoA models exhibited broader credible intervals and tended to overshoot the data. As a result, the UoM forecasts performed better than the ensemble forecasts (mean skill score of 0.31, see appendix S4).

Daily case incidence in Victoria steadily decreased from the August peak. In early September the Victorian government announced a gradual easing of restrictions, subject to reaching specific 14-day moving average case thresholds. Because the ensemble forecast comprised individual trajectories from each contributing model, we were able to calculate the proportion of all trajectories that satisfied a given 14-day case threshold on each date in the forecast horizon, and reported this as the daily probability of achieving each target threshold. These predictions were provided to the Victorian government throughout September and October.

### 3.5 Delta: June to December 2021

The onset of the Delta variant in June 2021 marked the first instance of sustained local COVID-19 transmission for most Australian jurisdictions, and public health responses focused on preventing local transmission. Accordingly, daily case counts remained very low in most jurisdictions, only exceeding 25 cases per day in the Australian Capital Territory (peak of 51), New South Wales (peak of 1,495), and Victoria (peak of 1,955). Ensemble forecasts for this period are presented in appendix S1.

Perhaps the single greatest value of the forecasts in this period was to demonstrate how rapidly case counts could increase if local transmission was left unchecked.

In May 2021, New South Wales Health set a target of achieving 80% coverage of the adult population with two vaccination doses by the end of December 2021 [31]. This was followed in August 2021 by an interim target of 70% coverage of the adult population with two doses, to encourage vaccine uptake and to begin easing restrictions for fully vaccinated people [31]. As a consequence, there was substantial vaccine roll-out over the 4-week forecast horizons from August onward. This was accompanied by rapid model development from the DST and UoM teams, with the effects of vaccination incorporated into these models in September 2021. The statistical MU model did not require such adjustments.

The forecasts were generally in very good agreement with the data in each jurisdiction. In particular, the mean CRPS values for New South Wales and Victoria were consistently lower than for the Pre-Delta wave in Victoria (see appendix S5).

### 3.6 Omicron BA.1: January to March 2022

This wave coincided with a national pivot from pursuing strong suppression to reopening and management of substantial levels of local COVID-19 transmission. Ensemble forecasts for this period are presented in appendix S1.

The forecasts struggled to predict the massive surge and subsequent decline in daily case counts, as demonstrated by decreased coverage (Figure 4), increased bias (Figure 5, appendix S3), and worse calibration than other waves (appendix S6).

This was due, at least in part, to mechanistic models failing to account for significant reduction in vaccine protection against Omicron, rapid changes in case ascertainment [32], and reduced mixing due to school holiday effects over December and January. Also in January, the vaccine roll-out began for children aged 5–11 years, and booster doses were recommended for at-risk individuals [33].

A primary concern in all jurisdictions was the timing of the epidemic peak, because the substantial increase in cases was impacting workforce capacity and causing disruptions to food supply chains, which in turn prompted panic buying [34]. The forecasts consistently predicted that the peaks would occur around 2 weeks later than they actually occurred (and for the mechanistic models, this was robust to adjustments such as relaxing assumptions regarding case ascertainment and immunity). Despite this inaccuracy, the forecast predictions that case activity would peak and begin to decrease in a matter of weeks remained an important and useful message for government.

### 3.7 Omicron BA.2/BA.5: April to August 2022

This period saw the gradual replacement of Omicron BA.1 with Omicron BA.2 and BA.5, and case counts in all jurisdictions steadily decreased after the large Omicron BA.1 peaks. Ensemble forecasts for this period are presented in appendix S1.

The forecasts reported on 11 June, generated from daily case counts up to 31 May (inclusive), predicted downwards trends in all jurisdictions, consistent with the trends in the most recent case counts. On 16 June we received additional genomic data from New South Wales that allowed us to estimate the transmission advantage of Omicron BA.5, relative to Omicron BA.1 and BA.2. By incorporating this transmission advantage into the mechanistic models, our next forecasts predicted upwards trends in all jurisdictions *despite no such trend in the reported aggregate data*. Consistent with these predictions, daily case counts began to increase later in June, and all jurisdictions experienced a peak in July (see figure S4).

By predicting this inflection point before it occurred, and by doing so based on appropriate interpretation of relevant local data, these forecasts helped convince public health stakeholders that the current downwards trend in cases would not be sustained.

### 3.8 Multiple variants: September to December 2022

Following the peak and decline of Omicron BA.5, Australia experienced circulation of numerous COVID-19 variants, with no single variant becoming dominant. Data from

international contexts provided evidence that many of these variants had growth advantages over Omicron BA.1, BA.2, and BA5, although it was unclear how these estimates might translate to Australia’s immune landscape. Ensemble forecasts for this period are presented in appendix S1.

Relative to the preceding Omicron BA.2/BA.5 period, the forecasts exhibited similarly good coverage (Figure 4) and performance (appendix S8). When case counts began to increase in November, the forecasts were confident that the epidemic peaks would be similar in size, or smaller than, the Omicron BA.5 peaks. This was a reassuring message that would prove to be accurate, and brings us to the end of our study period.

### 3.9 Communication of outputs

Over the 3.5 years of work summarised here, an ongoing concern was to ensure that model predictions and other quantitative outputs were reported in such a way that our stakeholders (spanning state, territory, and national governments and public health committees) would be able to interpret them appropriately and accurately act on and communicate their implications. While we did not have the capacity to undertake formal assessments of how the 152 weekly situation reports that we produced over this study period were interpreted, as described in McCaw and Plank [35] we did have open communication channels with our stakeholders and were able to explore and refine our reporting and visualisations in an iterative manner.

Figure 8 shows several examples of visualisations included in these reports. For example, our primary form of communicating the forecasts was daily credible intervals (see, e.g., Figure 3). However, this form can obscure the predicted size and timing of an epidemic peak, and so we explored the use of density plots to extract these features from individual forecast trajectories (Figure 8a). Another simple, yet surprisingly useful, modification to the daily credible interval figures was to overlay several randomly-selected forecast trajectories (Figure 8b), which helped to avoid the credible interval contours being mis-interpreted as case count trajectories. Finally, plotting past forecasts against the most recent data (Figure 8c) was a simple way of communicating recent forecast performance that provided stakeholders with a qualitative basis for deciding how much trust to place in the current forecasts.

**Figure 8:**
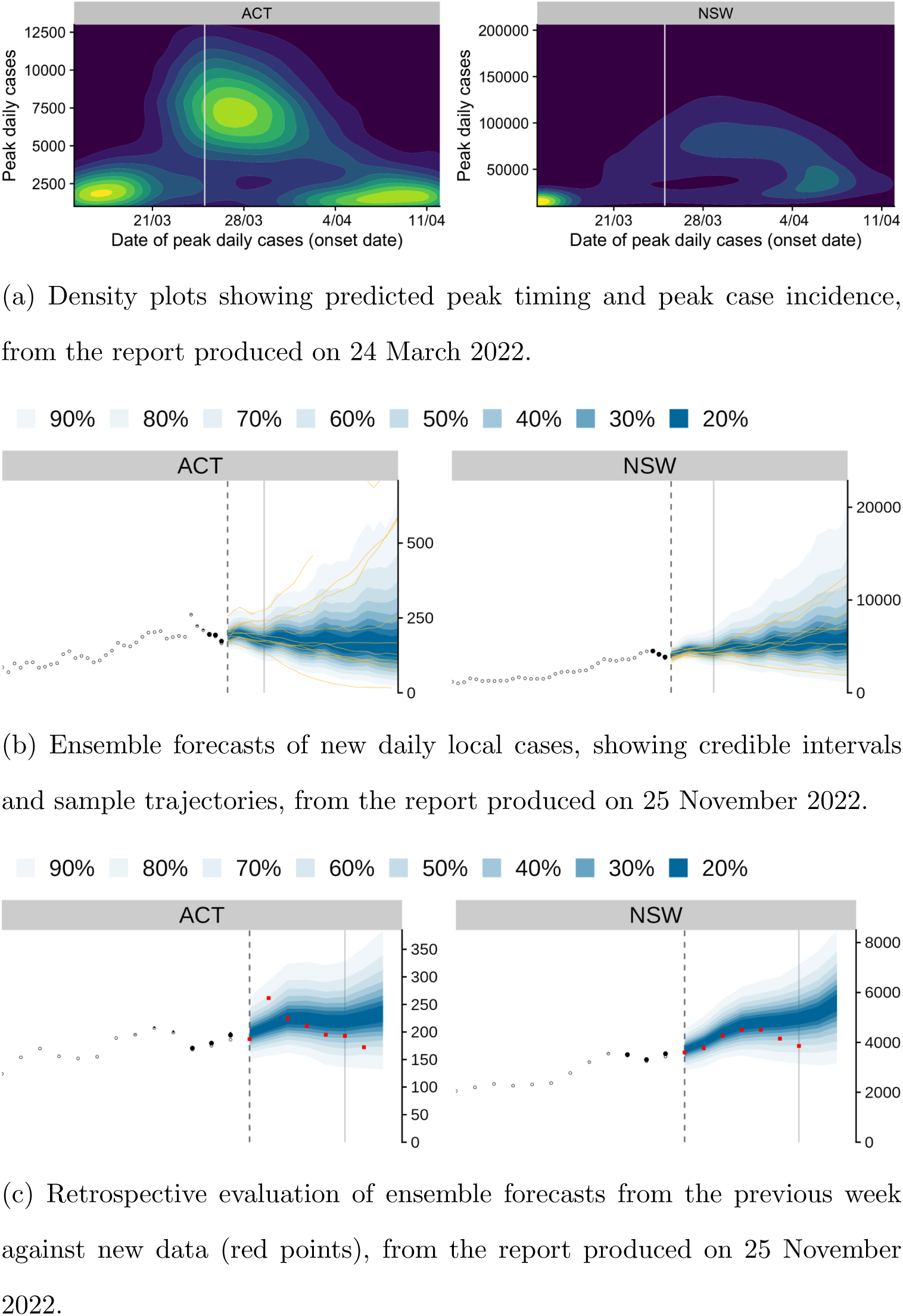
Example figures from weekly situation reports.

## 4 Discussion

### 4.1 Principal findings

Similar to findings reported in COVID-19 forecasting studies in other countries [6–11, 28, 29], the use of an ensemble to combine forecasts from multiple models improved forecast performance and reliability (Figures 4–6). Our ensemble forecasts provided information that supported public health responses throughout the study period, from the early periods of preventing local transmission, to the later periods of sustained local transmission and concerns about the burden placed on healthcare systems. Notable outcomes during the study period include:

- Pre-Delta: we used the forecasts to predict when 14-day average case targets would be achieved, which would trigger the gradual easing of mobility restrictions in the state of Victoria.
- Delta: this was the first instance of sustained local transmission for most jurisdictions, and the forecasts were useful for illustrating how rapidly case counts could increase if transmission was not curtailed.
- Omicron BA.1: the peak occurred 1–2 weeks earlier than the forecasts predicted, but forecast predictions that activity would peak and then decline in a matter of weeks was reassuring in the face of workplace absenteeism that disrupted supply chains.
- Omicron BA.2/BA5: the forecasts were able to correctly predict an increase in cases while the current case counts were decreasing, and this was a particularly useful message for health protection committees.
- Multiple variants: the forecasts accurately predicted that the peaks in late 2022 would not be larger than the Omicron BA.5 peaks.

### 4.2 Study strengths

The ensemble approach allowed us to readily incorporate forecasts contributed by teams across Australia, and provided a framework for rapid evaluation of new model iterations for potential inclusion in the ensemble. The weekly performance evaluations and model inclusion/exclusion decisions relied on measures of forecast error (using CRPS) under the assumption that existing policies would persist for the entire forecast horizon. However, as we have described above, the ensemble forecasts influenced policy decisions throughout the study period, and some of these decisions directly impacted local COVID-19 transmission and/or case ascertainment. Accordingly, when policy decisions were understood to have influenced the case data reported over a forecast horizon, we factored these effects into our analyses. In particular, where the difference between forecasts and reported data were consistent with the likely effects of the policy decisions, we considered this a successful outcome [36].

We deliberately chose to use equal-weight ensemble forecasts and, as we explain in the methods section, each model was evaluated against the most recent data rather than competing against the other models for inclusion in the ensemble [26]. As we have discussed in the results section, a model’s ranking in recent weeks was not a reliable predictor of that model’s ranking in future weeks. The strongest finding we have regarding the rankings is that the ensemble was either the best, or second-best, more often than any individual model. Both findings are consistent with ensemble COVID-19 forecast evaluations in other countries [9, 10].

Finally, we note here that the ensemble forecasts presented here also served as an input for COVID-19 hospital bed occupancy forecasts (December 2021 to December 2023) that further supported Australia’s public health responses [18].

### 4.3 Study limitations

While pursuing the prevention of local transmission, Australian jurisdictions maintained high testing levels and the proportion of infected persons that were identified as cases was likely to be both very high, and to remain relatively constant. The ensemble forecasts performed very well during this period (the “Pre-Delta” and “Delta” phases).

When Australia transitioned to re-opening, case ascertainment was substantially reduced, and was challenging to estimate in near-real-time [32]. By February 2022, more than 94% of people over the age of 16 were fully vaccinated [37], and while the mechanistic models were subject to numerous adaptations and refinements to account for vaccination coverage, booster vaccinations, and waning immunity, throughout 2022 it became more challenging to estimate population susceptibility to the emerging variants of concern. This was due to an increasingly complex population immune landscape, resulting from heterogeneous levels of vaccination and immunising exposure, and challenges in estimating the immunogenicity of emerging variants.

However, despite these challenges, the inclusion of both mechanistic and statistical models in the ensemble substantially improved the forecast performance and reliability across the entire study period. The only period where the forecast did not capture the case data was the rapid emergence and decline of Omicron BA.1, which occurred as Australia pivoted from pursuing strong suppression to reopening and accepting substantial levels of local transmission — a transition that was inherently challenging to predict.

### 4.4 Meaning and implications

Our ensemble COVID-19 forecasts were produced under contract with the Australian Government Department of Health and Aged Care, and participation was limited to a small number of institutes. In contrast, the United States of America and Europe both established public COVID-19 forecast hubs that had open submission policies and included all submissions that complied with the hubs’ technical requirements in their ensemble forecasts [9, 10]. This open nature resulted in large numbers of participating teams, with 67 teams contributing US forecasts (July 2020 to December 2021 [10]), and 48 teams contributing European forecasts (March 2021 to March 2022 [9]). Both hubs reported that this open nature also limited the possibility to understand drivers of forecast performance, with many teams participating at different times, participating intermittently, and providing varied and/or limited descriptions of their methods [9, 10].

Forecast evaluations from both hubs reported findings that are broadly consistent with those reported here. Forecasts performed well in periods of stable behaviour, but struggled at longer horizons around inflection points, and individual models varied widely in their ability to account for new COVID-19 variants. The ensemble forecasts were consistently among the best-performing forecasts across all horizons and locations, and weighting individual models by their past performance did not improve the ensemble performance. The US hub reported that forecast prediction intervals were generally over-confident and had low coverage, particularly when case numbers were changing rapidly [10], similar to our findings for the Omicron BA.1 wave (Figure 4).

The ensemble forecasts presented here played an important role in supporting Australian public health decision-making over a wide range of epidemiological and policy contexts. Consistent with reflections from modelling and data analysis communities around the world [35, 38–40], frequent communication between modellers and public health stake-holders — and the mutual understanding and trust that this fostered — was integral to the utility of these ensemble forecasts. Being organised as a consortium, with several members sitting on key public health committees, meant that there were direct lines of communication between decision makers and modellers, and this in turn expedited the prototyping and development of effective analyses and communications.

More broadly, the collective role of our consortium as a means for weekly peer-review of model forecasts, and the weekly decision of which model iteration(s) to include, directly supported rapid model development and evaluation, while also ensuring that only “known good” model iterations were contributing to the ensemble forecast. Having common performance targets and evaluation processes for all models, conducted openly within the consortium each week, fostered mutual collaboration. This collaborative structure also helped to avoid duplication of effort, which was extremely beneficial at times of high stress, urgent delivery schedules, and unsustainable workloads. Such an approach may be challenging to adapt to the scale of the US and European forecast hubs.

A final benefit of this organisation, and the small size of our consortium relative to similar groups overseas, was retaining the human element in selecting which model itera-tion(s) to include in the ensemble, rather than using an arbitrary quantitative threshold or simply including all available forecasts in the ensemble. This allowed us to explain features of each ensemble forecast, and the underlying rationale for these features in terms of the contributing models and their assumptions, which helped our public health stakeholders to interpret the forecasts appropriately.

## Supporting information

Technical appendix

## Data Availability

The analysis code, summary outputs, figures, and supplementary materials are provided in a public git repository: https://gitlab.unimelb.edu.au/rgmoss/analysis-australian-covid19-ensemble-forecasts-results.
Note that this repository does not include the input cases files and output ensemble forecasts (approximately 2.3 Gb). These are provided in a separate data repository: https://doi.org/10.26188/29434298.

https://doi.org/10.26188/29434298

## 5 Ethics statement

The study was undertaken as urgent public health action to support Australia’s COVID-19 pandemic response. The study obtained data under the National Health Security Agreement for the purposes of national communicable disease surveillance. Contractual obligations established strict data protection protocols, as agreed between the University of Melbourne and sub-contractors and the Australian Government Department of Health and Aged Care. Oversight and approval for use in supporting Australia’s pandemic response, and for publication, were provided by the data custodians (represented by the Communicable Diseases Network of Australia, CDNA). The ethics of the use of these data for these purposes, including publication, was agreed by the Department of Health and Aged Care with CDNA. All methods were carried out in accordance with the relevant guidelines and regulations.

