## Supplementary material for "Ensemble forecasts of COVID-19 activity to support Australia’s pandemic response: 2020–22": Technical appendix

#### Contents

**S1 Ensemble forecasts for each phase of the study period** 2

**S2 Model rankings for each ensemble forecast** 7

**S3 Probability integral transform (PIT) histograms** 8

**S4 Model skill scores for the Pre-Delta wave in Victoria** 11

**S5 Forecast evaluations around observed epidemic peaks** 12

**S6 Marginal quantile and CDF calibration plots** 16

**S7 Daily case counts and CRPS values for each jurisdiction** 20

**S8 CRPS values for each dominant strain** 28

### S1 Ensemble forecasts for each phase of the study period

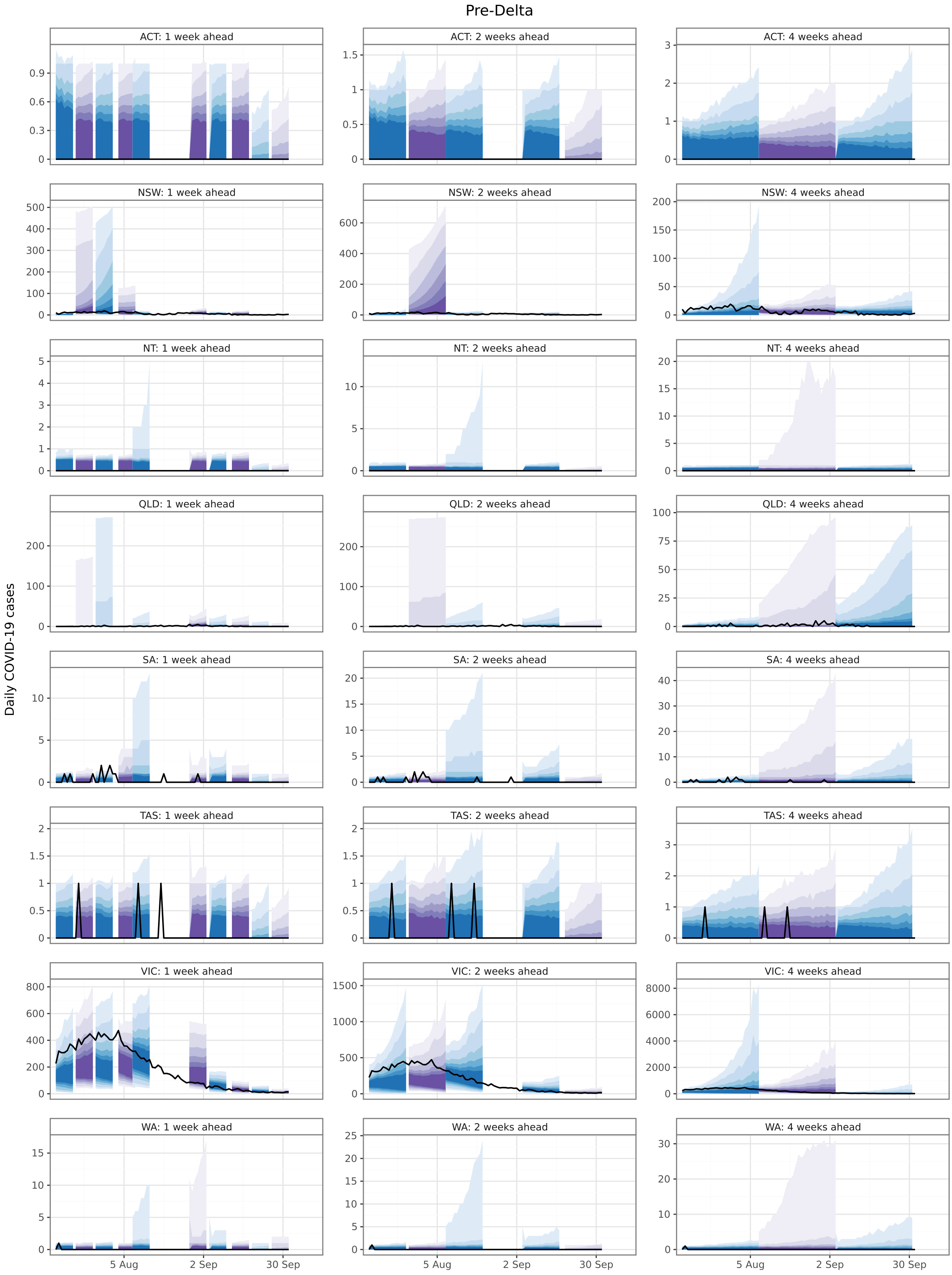


Figure S1: Ensemble forecasts for the "Pre-Delta" period (2020).

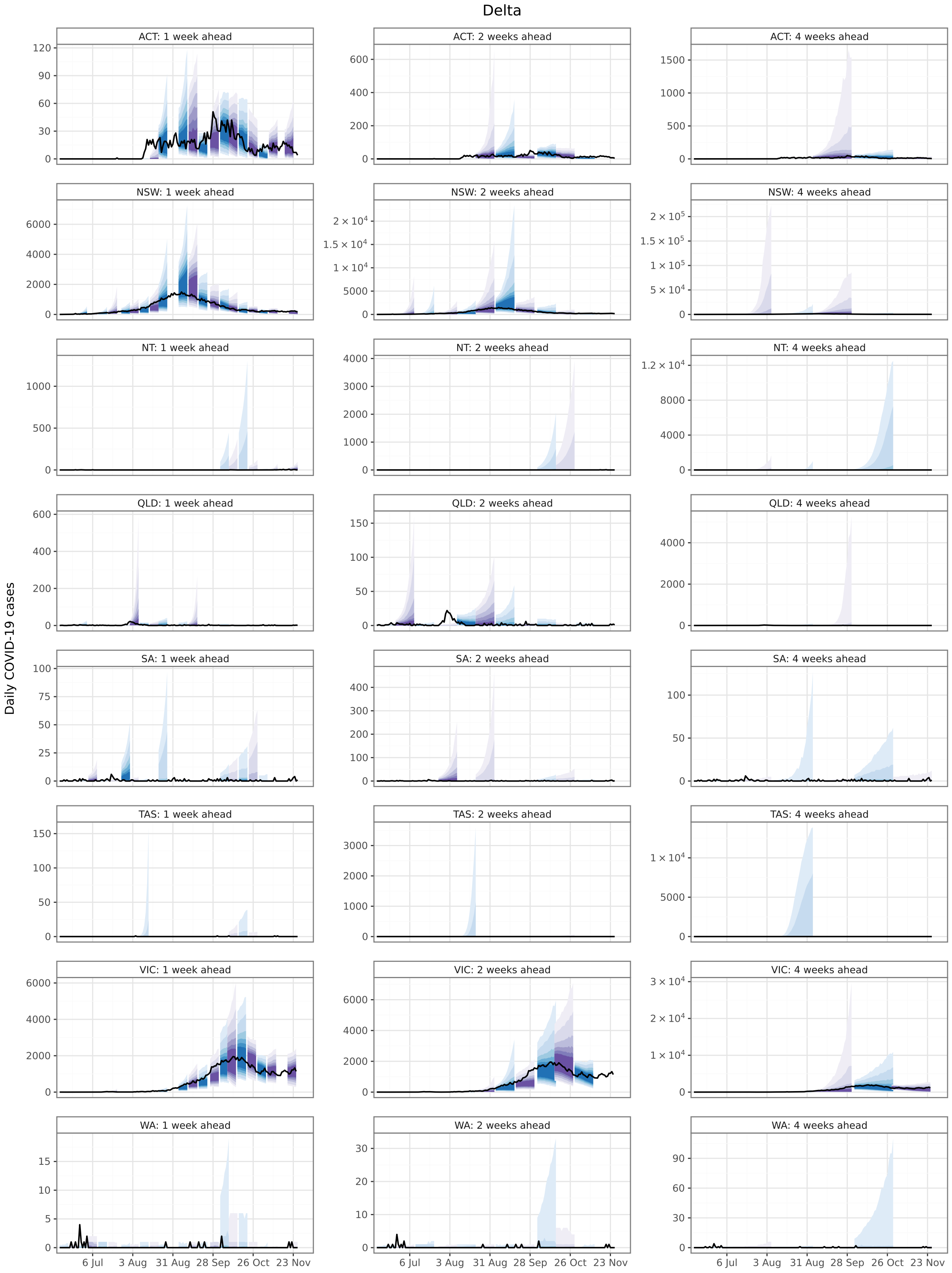


Figure S2: Ensemble forecasts for the “Delta” period (2021).

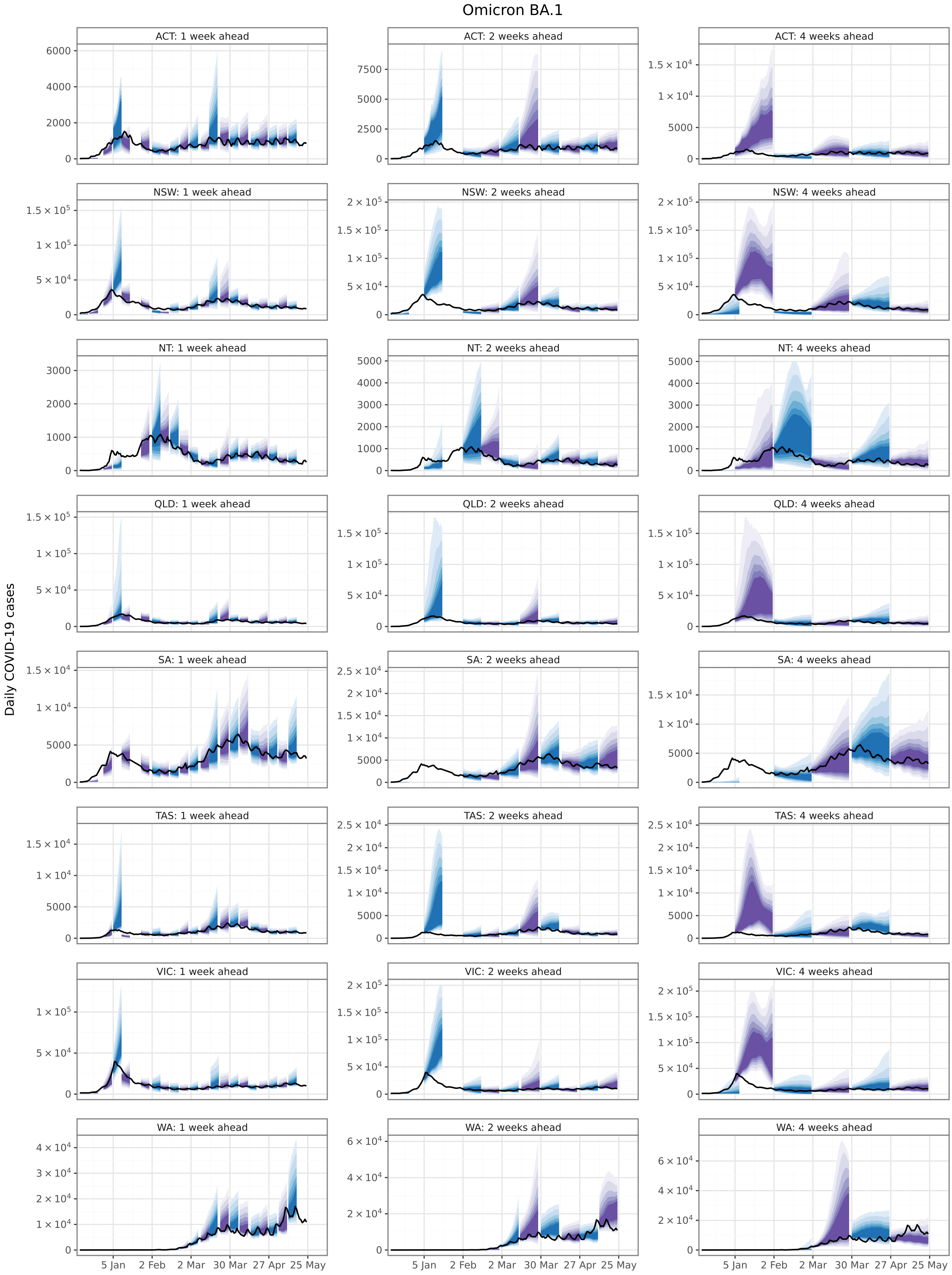


Figure S3: Ensemble forecasts for the “Omicron BA.1” period (2022).

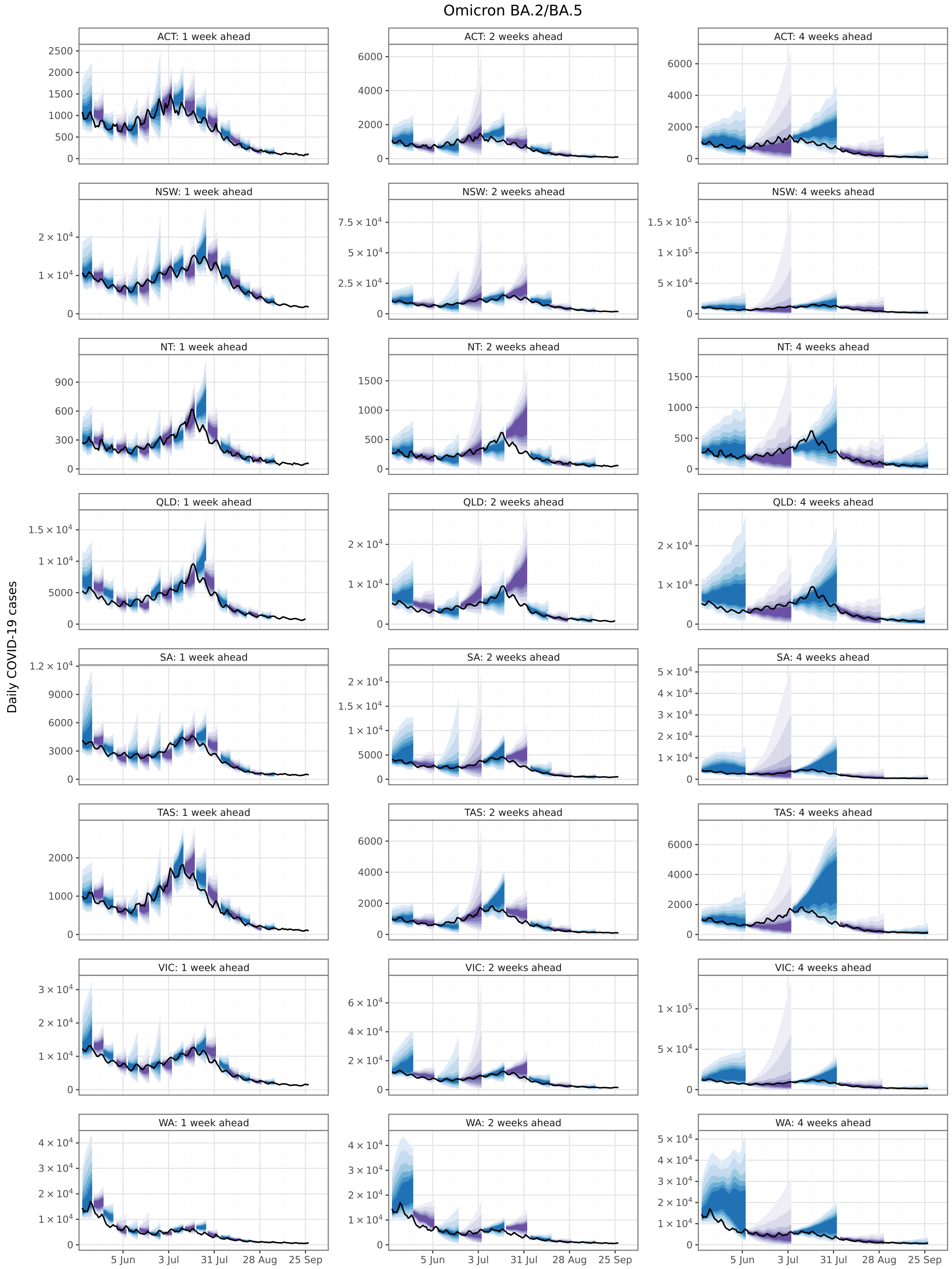


Figure S4: Ensemble forecasts for the "Omicron BA.2/BA.5" period (2022).

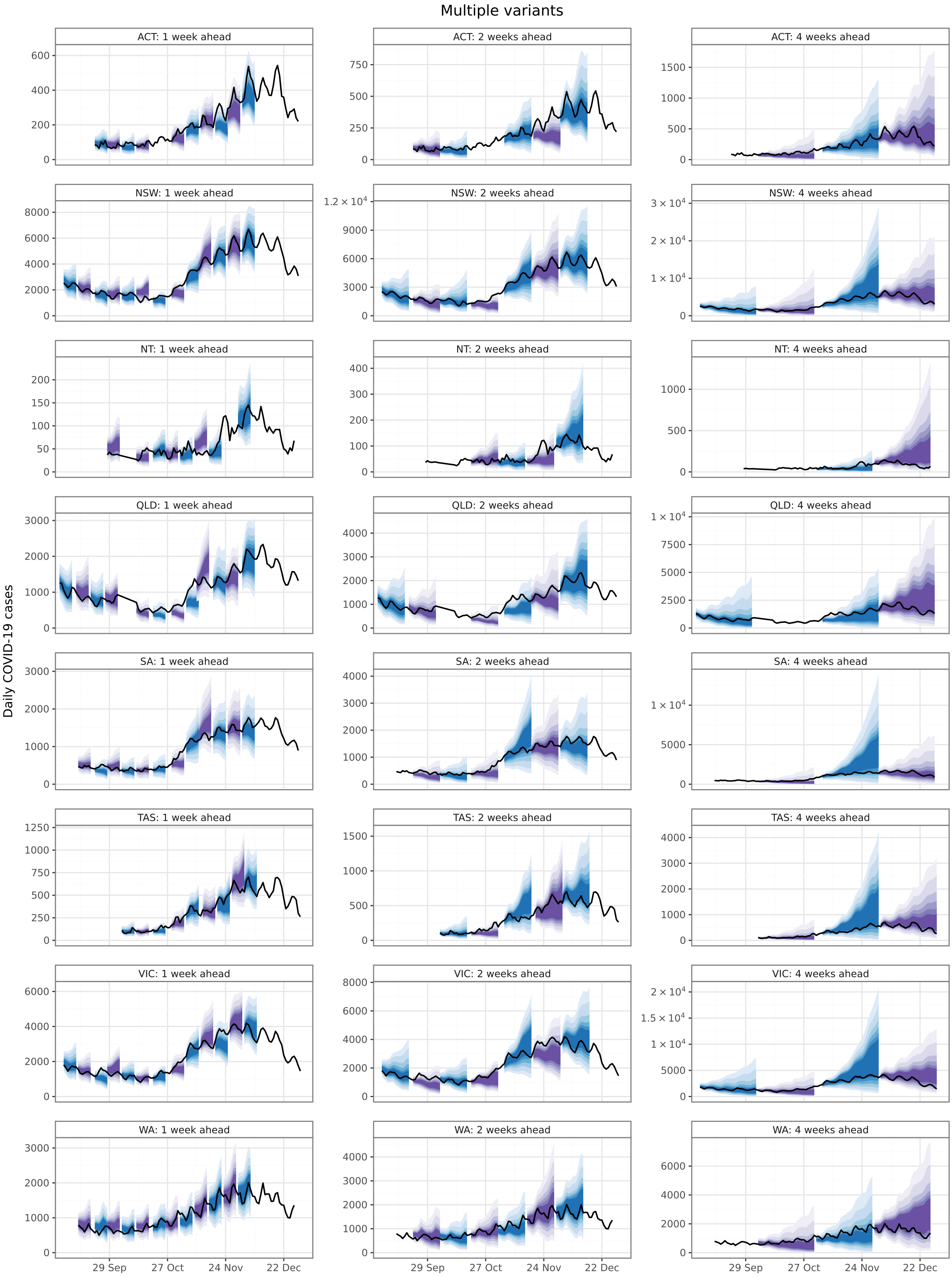


Figure S5: Ensemble forecasts for the "Multiple variants" period (2022).

### S2 Model rankings for each ensemble forecast

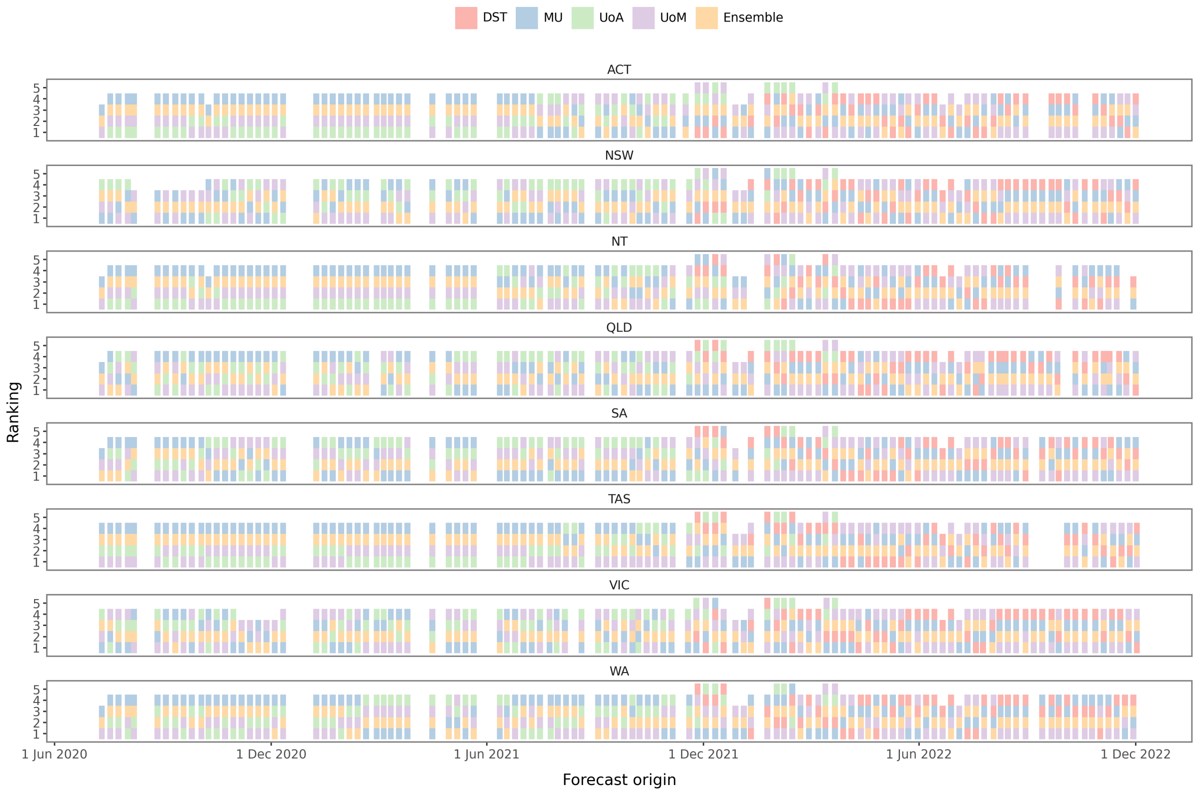


Figure S6: The model forecast rankings for each ensemble forecast.

#### S3 Probability integral transform (PIT) histograms

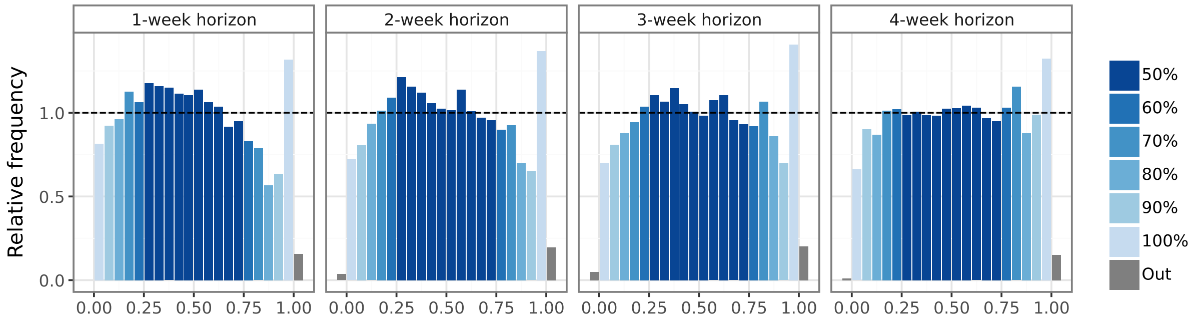


Figure S7: Probability integral transform (PIT) histograms showing ensemble forecast coverage for lead times of 1–7 days, 8–14 days, 15–21 days, and 22–28 days.

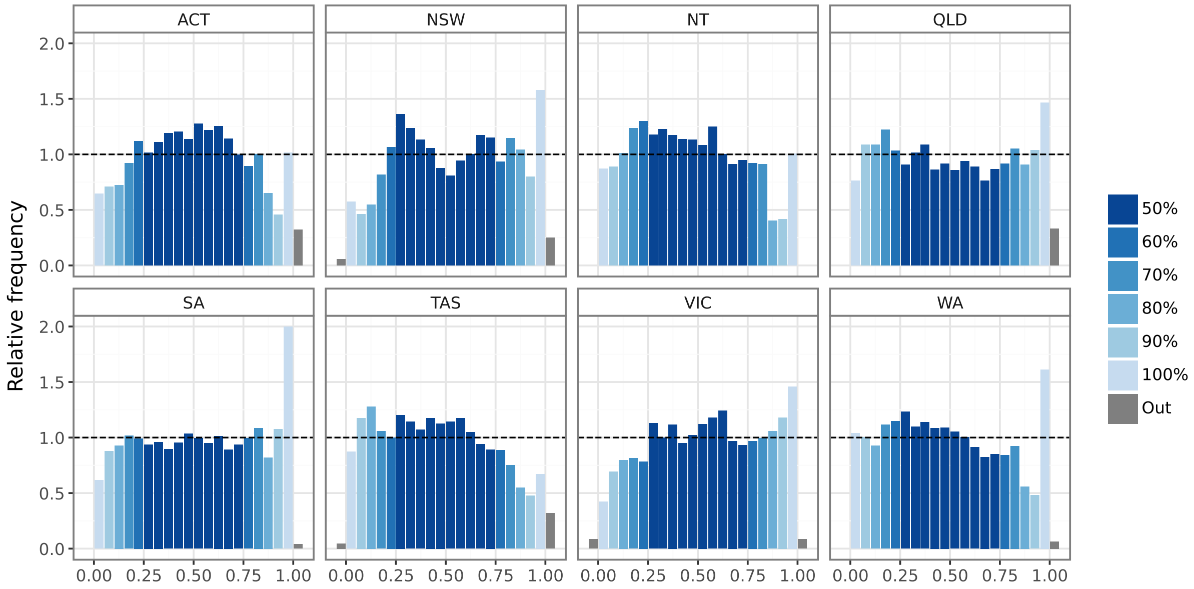


Figure S8: Probability integral transform (PIT) histograms showing ensemble forecast coverage for each jurisdiction.

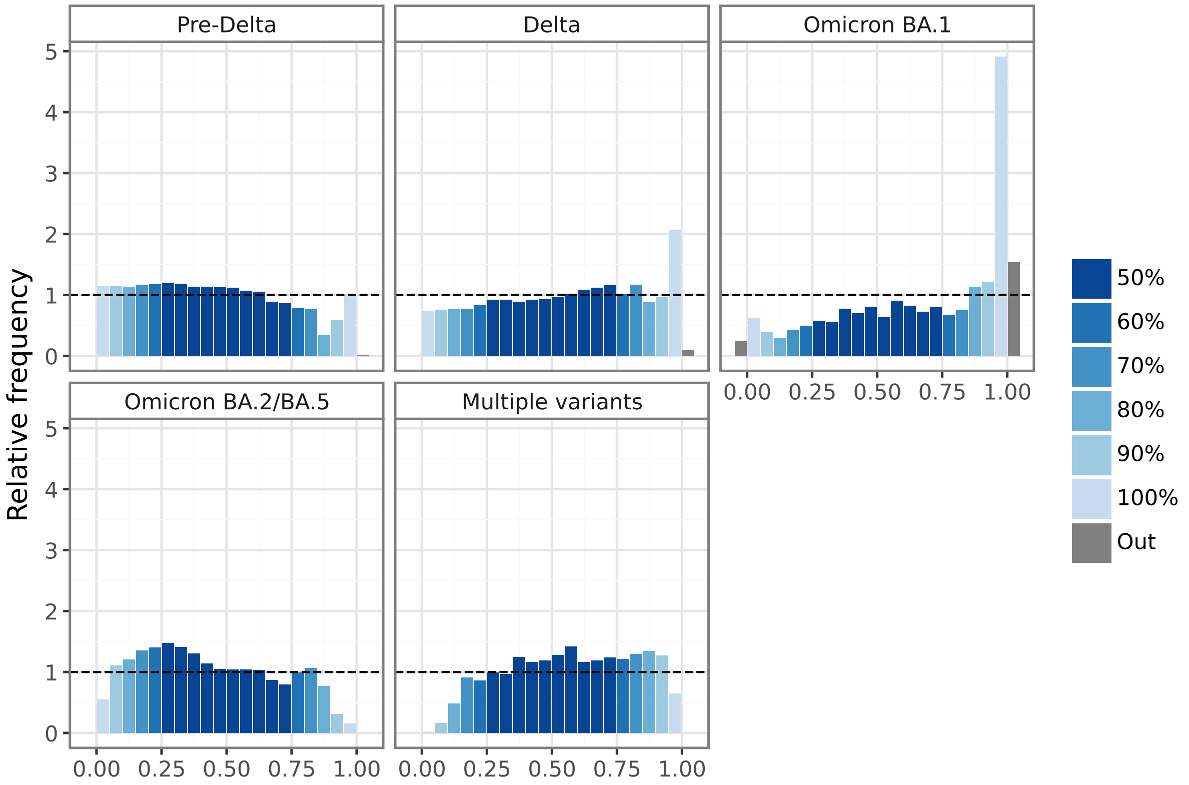


Figure S9: Probability integral transform (PIT) histograms showing ensemble forecast coverage for each period where disease activity was dominated by a particular strain or group of strains.

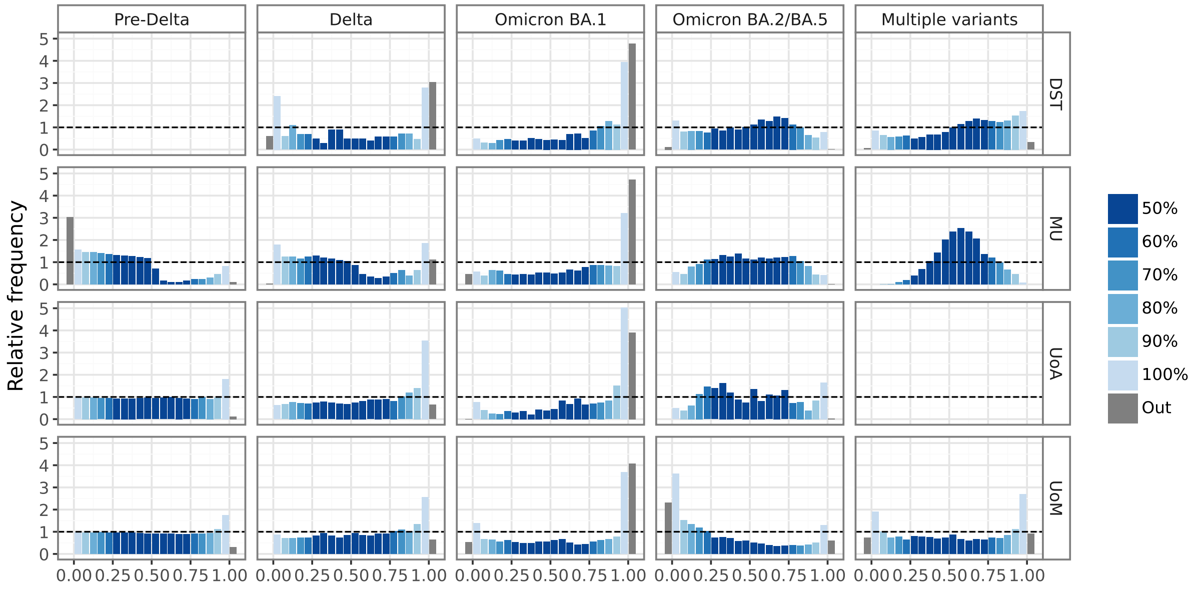


Figure S10: Probability integral transform (PIT) histograms showing forecast coverage separately for each model in the ensemble, for each period where disease activity was dominated by a particular strain or group of strains.

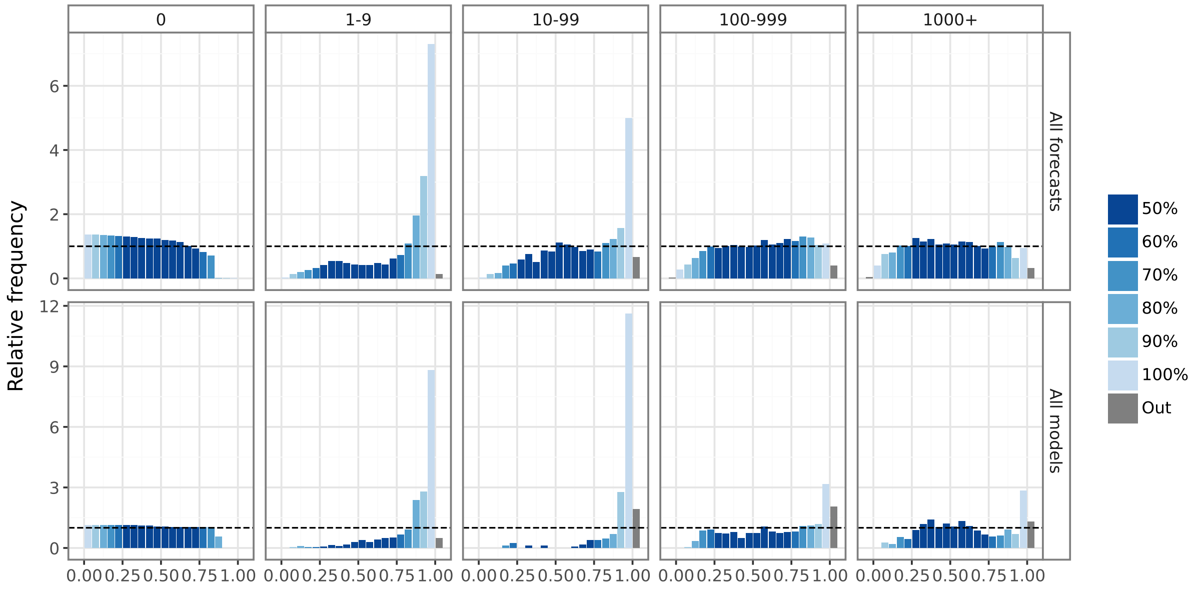


Figure S11: Probability integral transform (PIT) histograms showing ensemble forecast coverage for the whole study period (top row, “All forecasts”), and the interval where all models contributed to the ensemble (“All models”, bottom row).

#### S4 Model skill scores for the Pre-Delta wave in Victoria

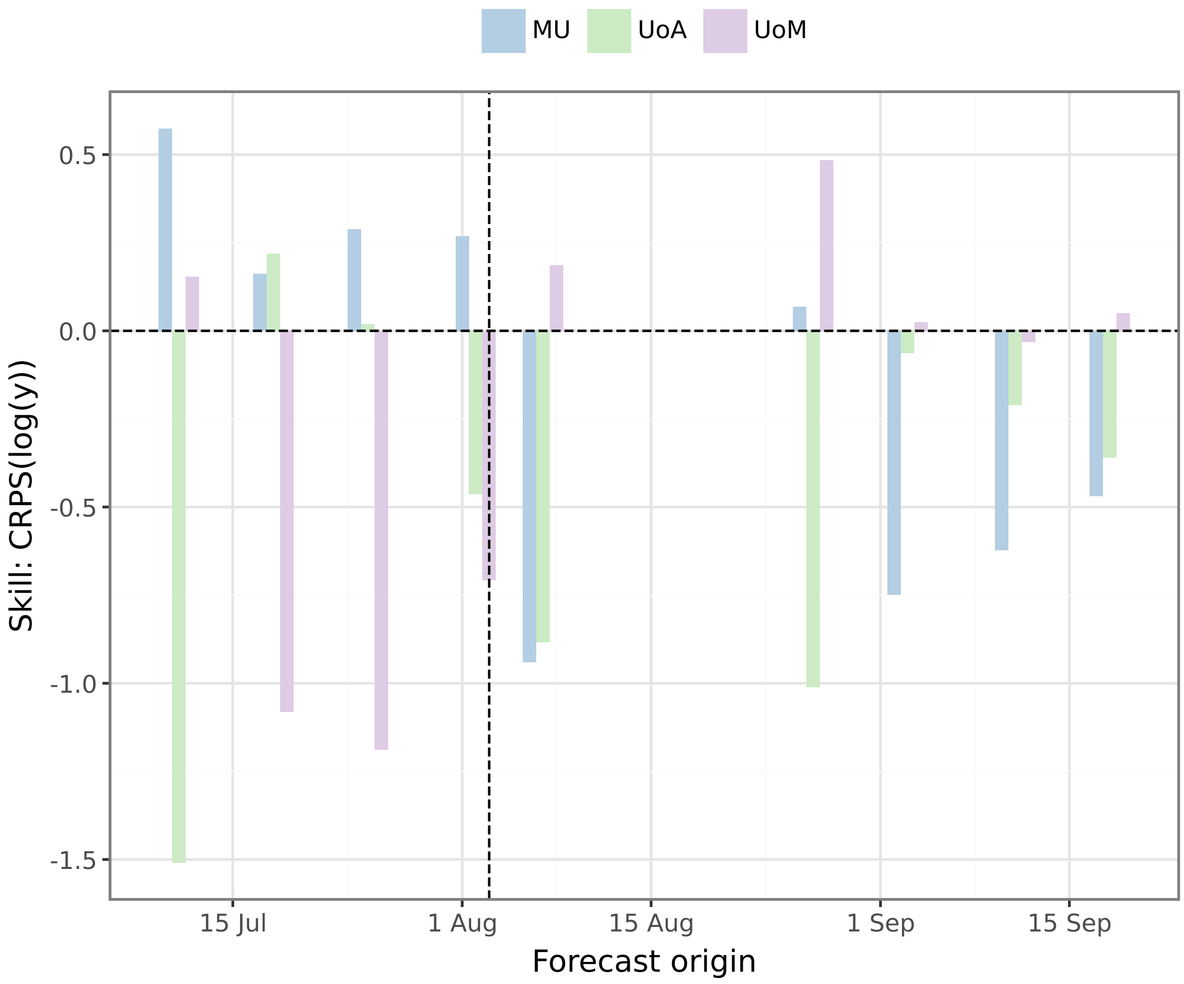


Figure S12: Model skill scores during the second wave in Victoria (May to October 2020) relative to the ensemble forecast. The true peak occurred on 3 August 2020, as indicated by the vertical dashed line. Skill scores were calculated using CRPS on log-transformed values.

#### S5 Forecast evaluations around observed epidemic peaks

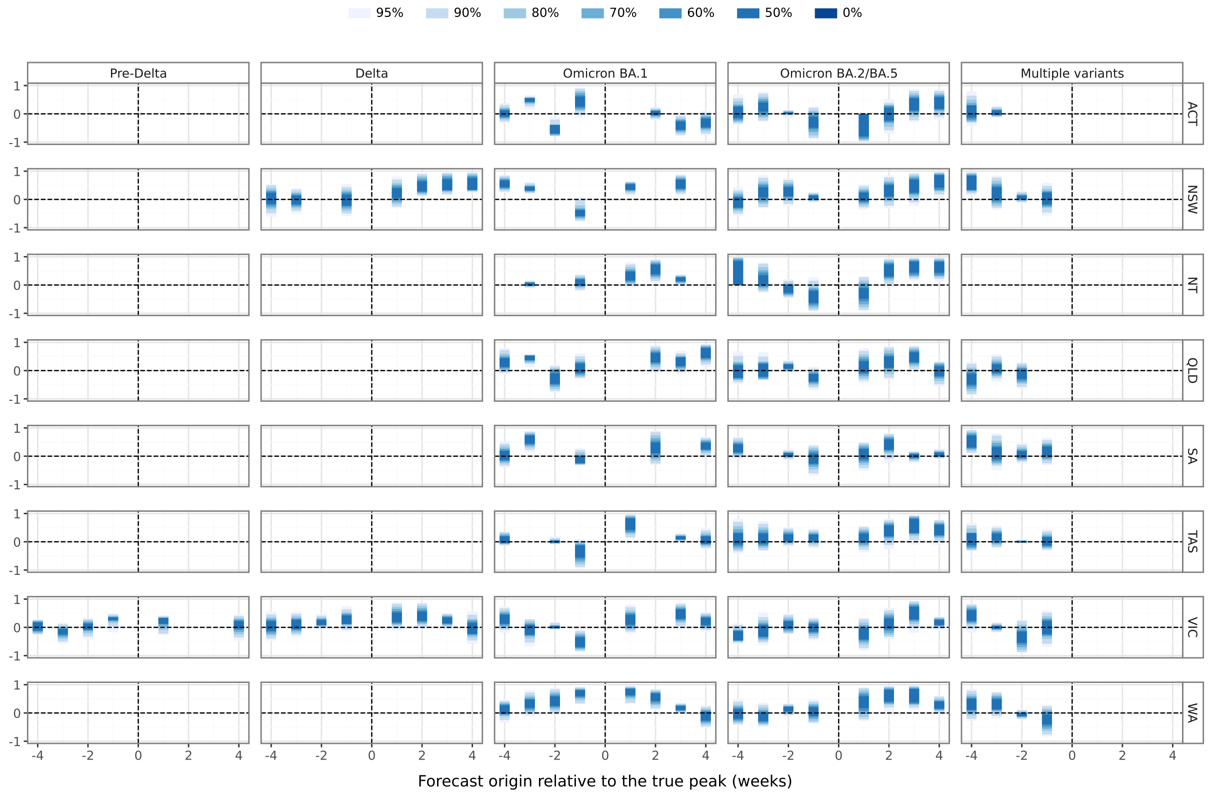


Figure S13: The correlation between forecast trajectories and the ground truth case counts, calculated for forecasts where the true peak occurred within the four-week forecast horizon (negative x-axis values) or the four weeks prior to the forecast (positive x-axis values).

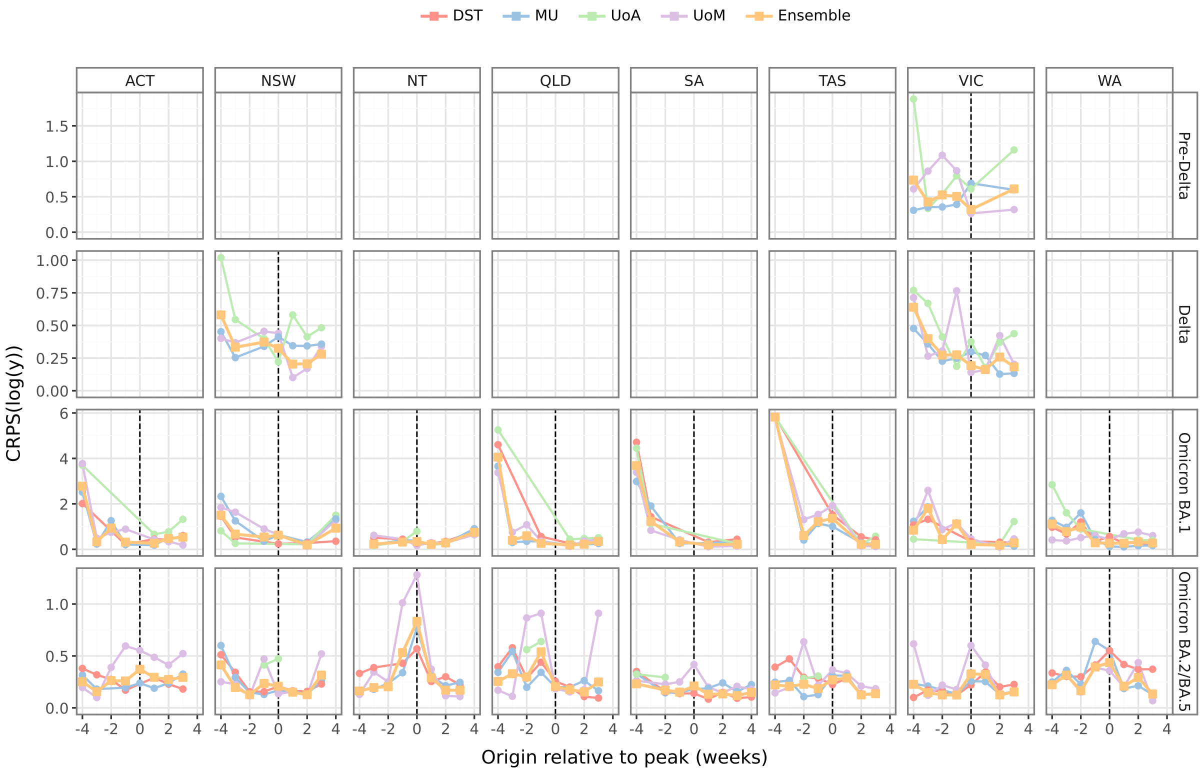


Figure S14: Mean forecast CRPS for each model and the ensemble, shown for forecast origins within 4 weeks either side of the largest peak observed for each dominant strain (minimum 200 cases), with the exception of the "Multiple variants" period where activity was flatter and observed peaks occurred towards the very end of the study period.

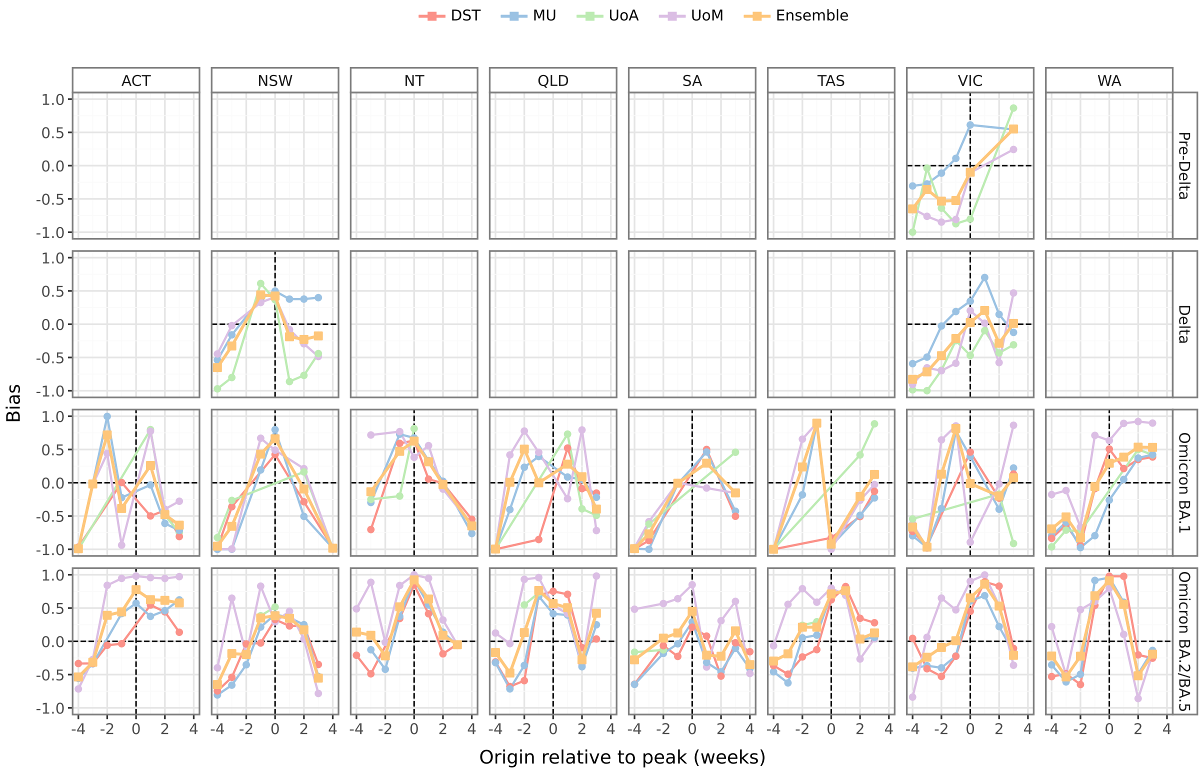


Figure S15: Forecast bias for each model and the ensemble, shown for forecast origins within 4 weeks either side of the largest peak observed for each dominant strain (minimum 200 cases), with the exception of the "Multiple variants" period where activity was flatter and observed peaks occurred towards the very end of the study period.

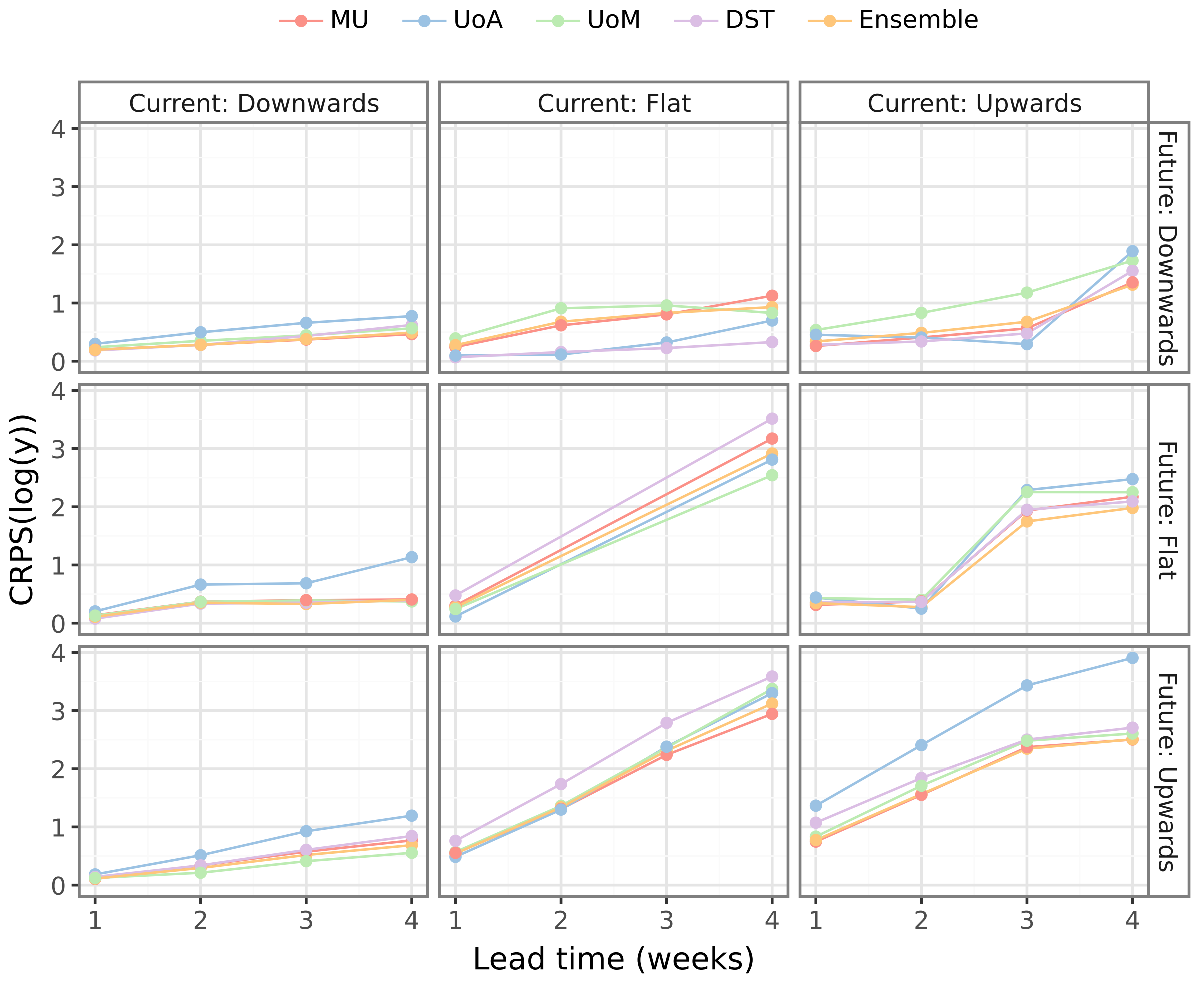


Figure S16: Mean CRPS values for predictions made within 5 weeks before/after the peak observed in each jurisdiction for Omicron BA.1. Values are shown for each model, for each week of the forecast horizon, and categorised by the trend in the most recent case data (columns: downwards, flat, upwards) and the trend in the data over the corresponding week of the forecast horizon (rows: downwards, flat, upwards).

#### S6 Marginal quantile and CDF calibration plots

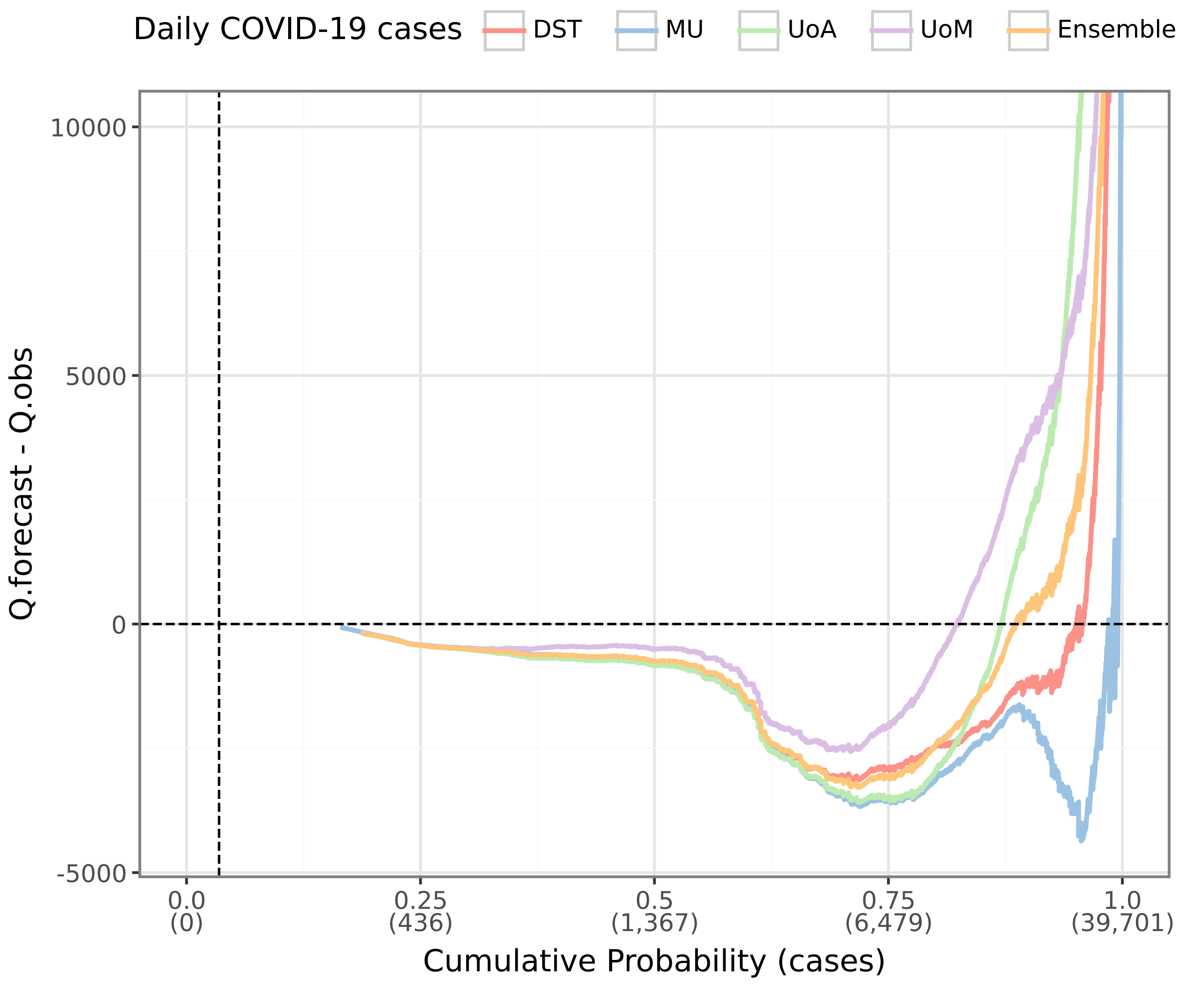


Figure S17: The marginal quantile calibration for each model, for the interval where all models contributed to the ensemble. Negative values indicate cumulative under-prediction and positive values indicate cumulative over-prediction. All models exhibited under-prediction between 1,000 and 6,000 daily cases, and over-prediction at the highest daily case counts ($\geq$ 30,000).

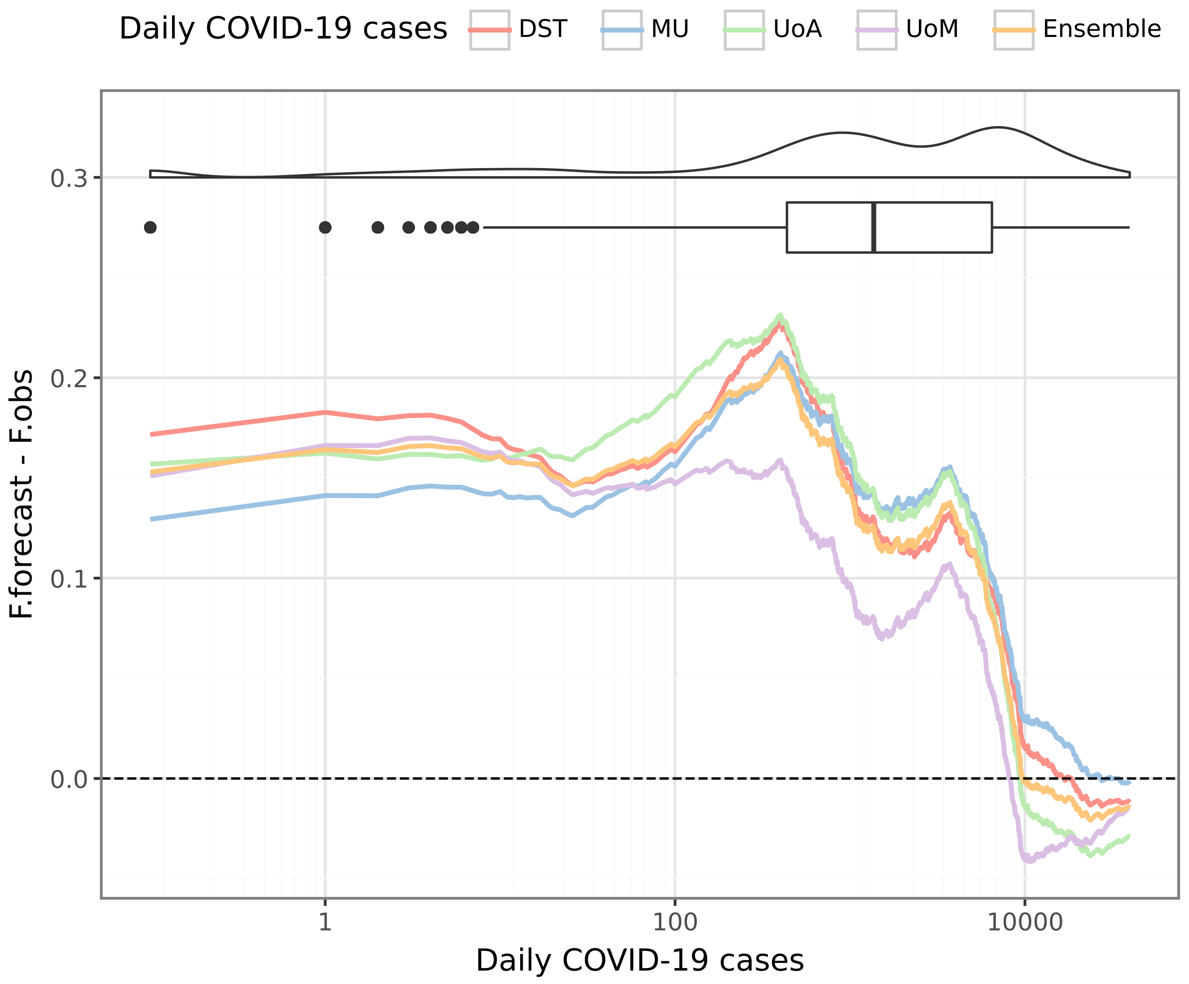


Figure S18: The marginal CDF calibration for each model, for the interval where all models contributed to the ensemble. All models over-predicted zeros and under-predicted the highest case counts. Both of these features primarily stem from the model performances during the Omicron BA.1 period (December 2021 to February 2022).

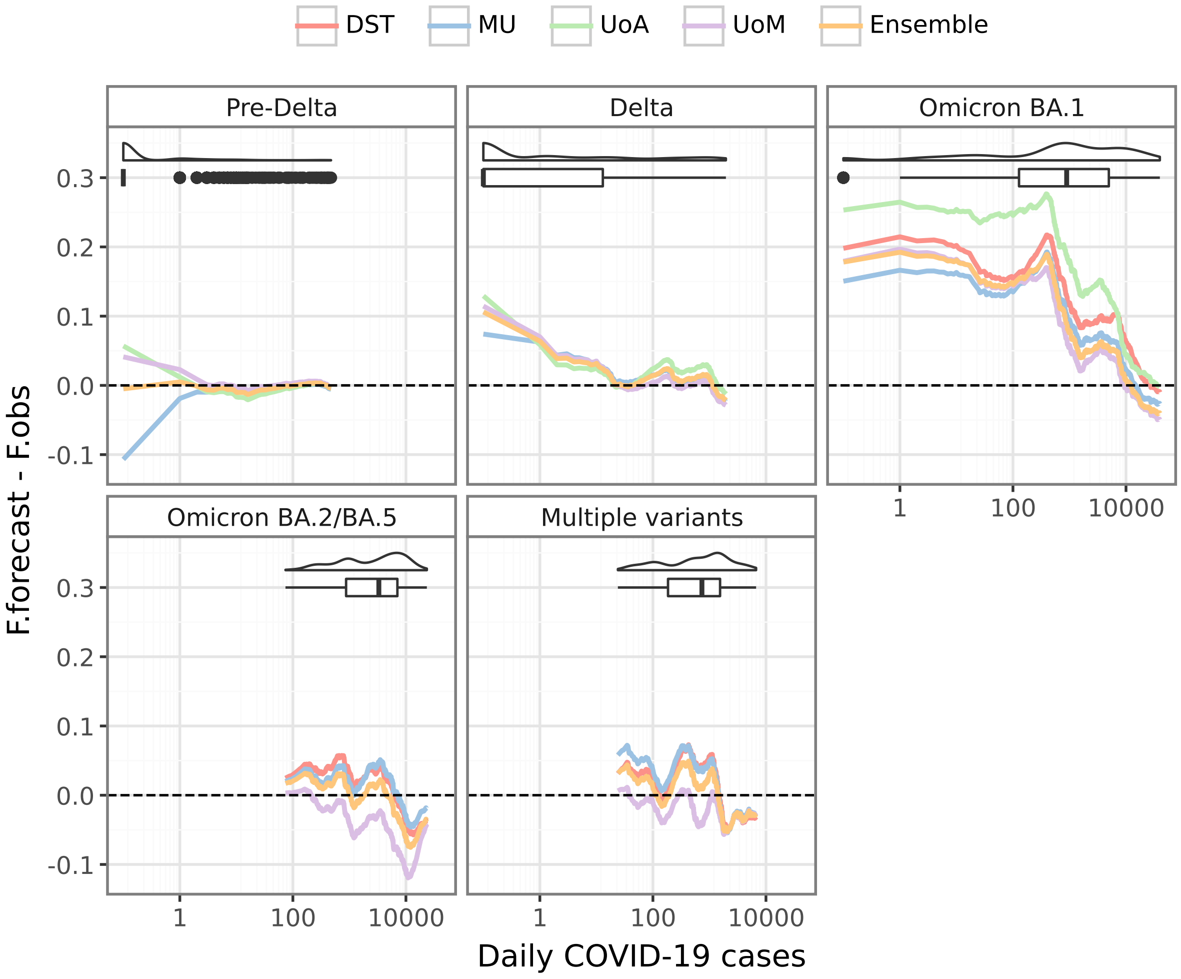


Figure S19: The marginal CDF calibration for each model, shown separately for each period where disease activity was dominated by a particular strain or group of strains. All models over-predicted zeros during the Delta and Omicron BA.1 periods, and under-predicted the highest case counts during the Omicron BA.1 period.

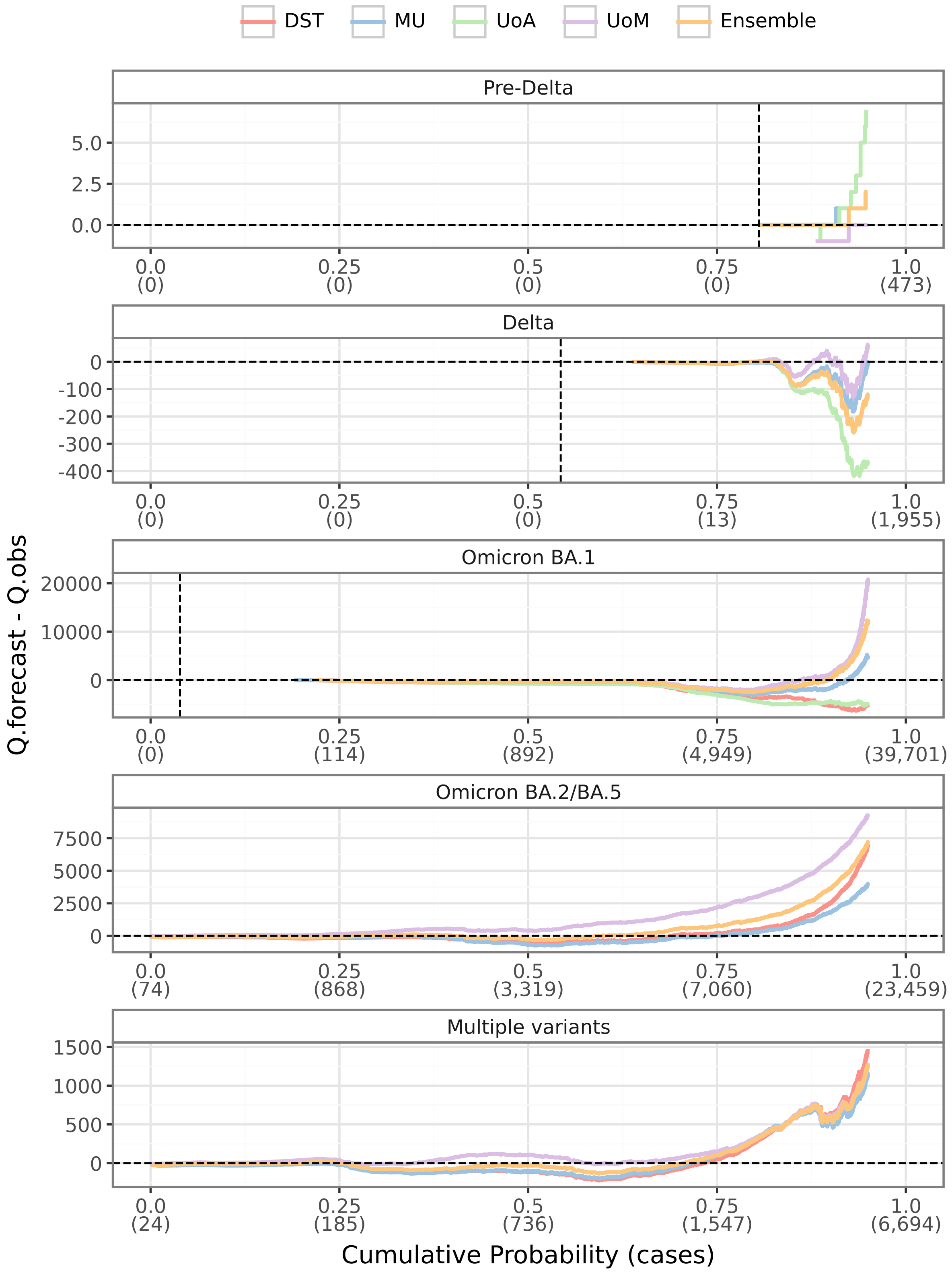


Figure S20: The marginal quantile calibration for each model, shown separately for each period where disease activity was dominated by a particular strain or group of strains.

#### S7 Daily case counts and CRPS values for each jurisdiction

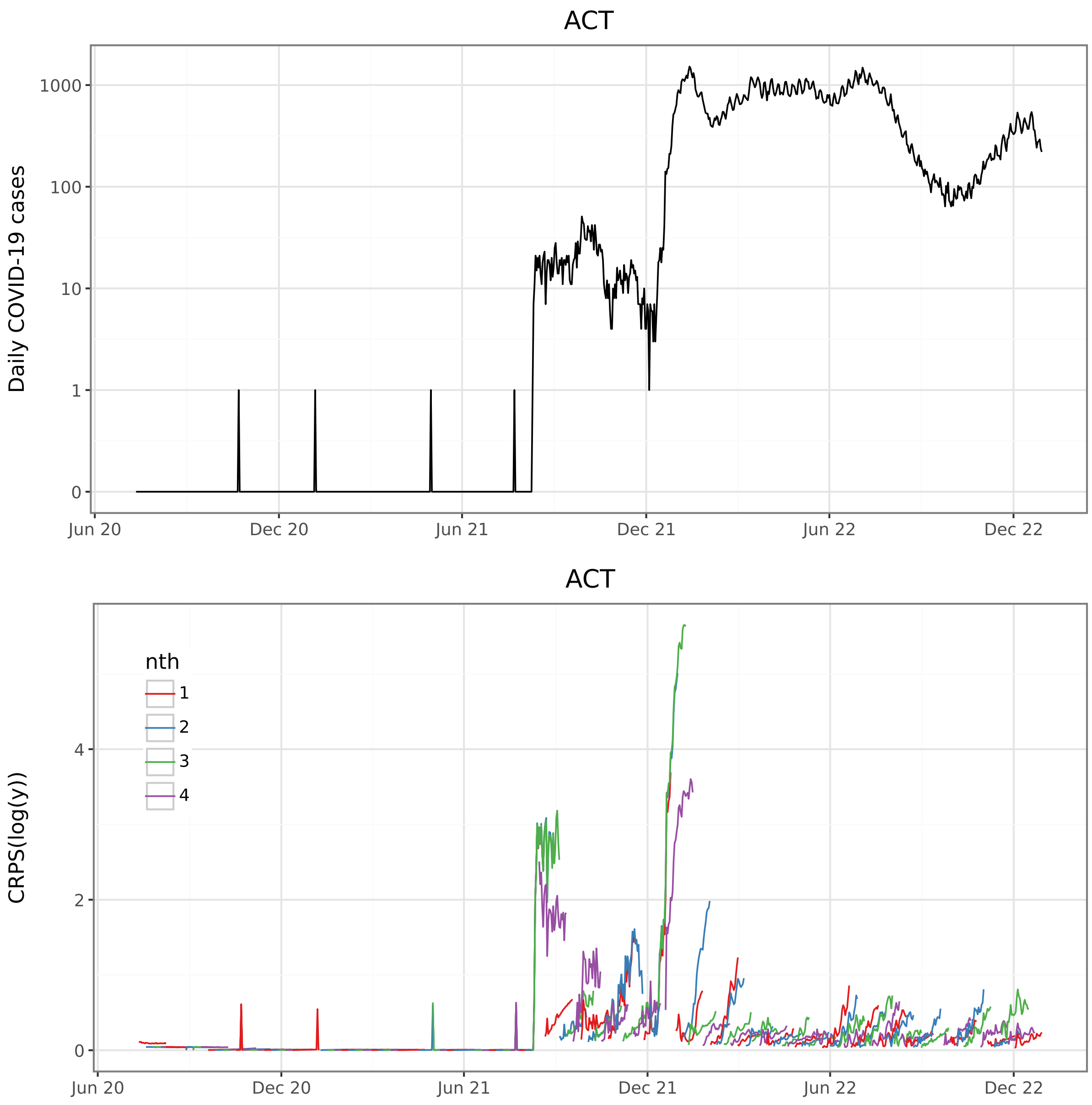


Figure S21: ACT daily cases and CRPS.

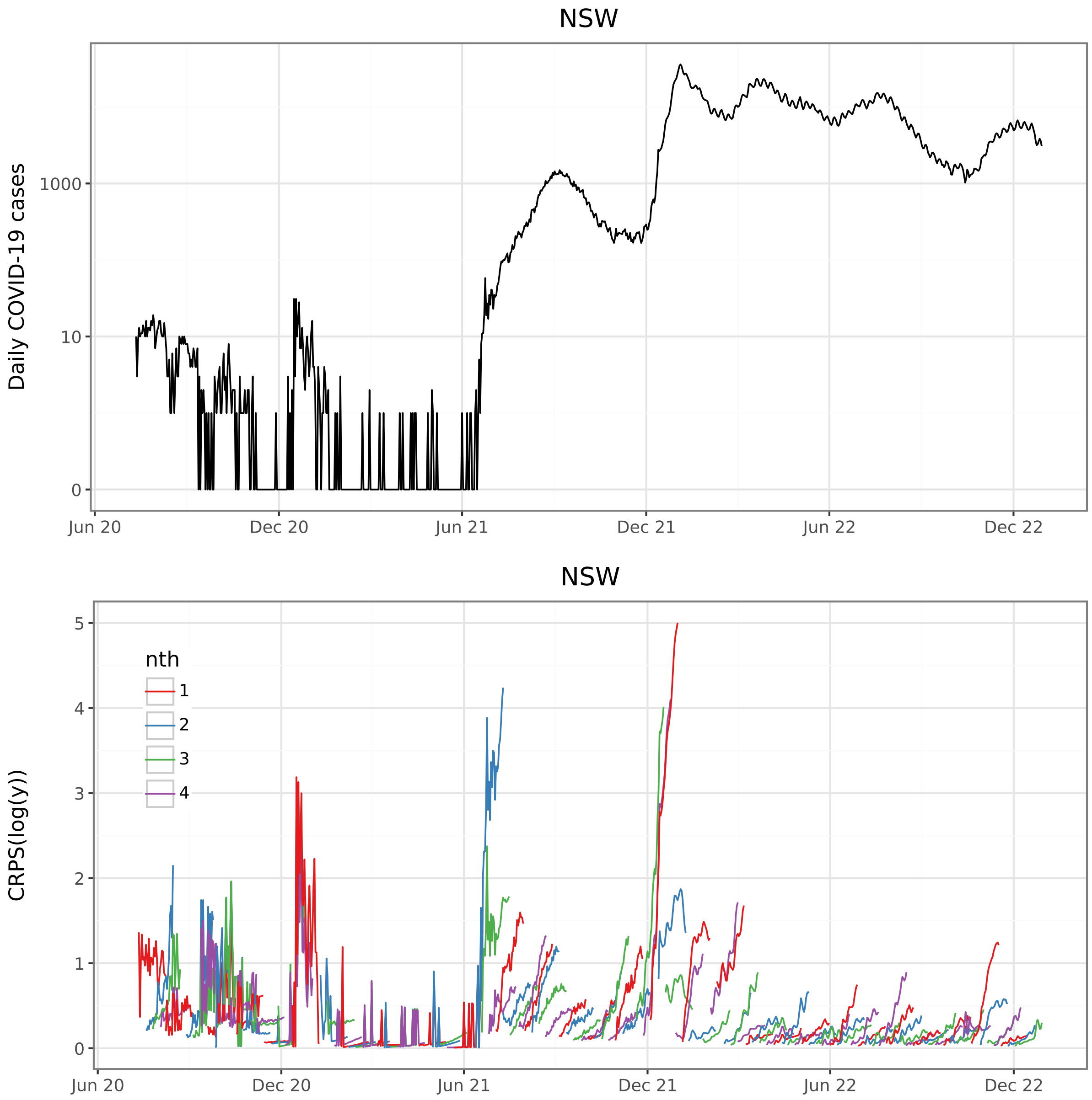


Figure S22: NSW daily cases and CRPS.

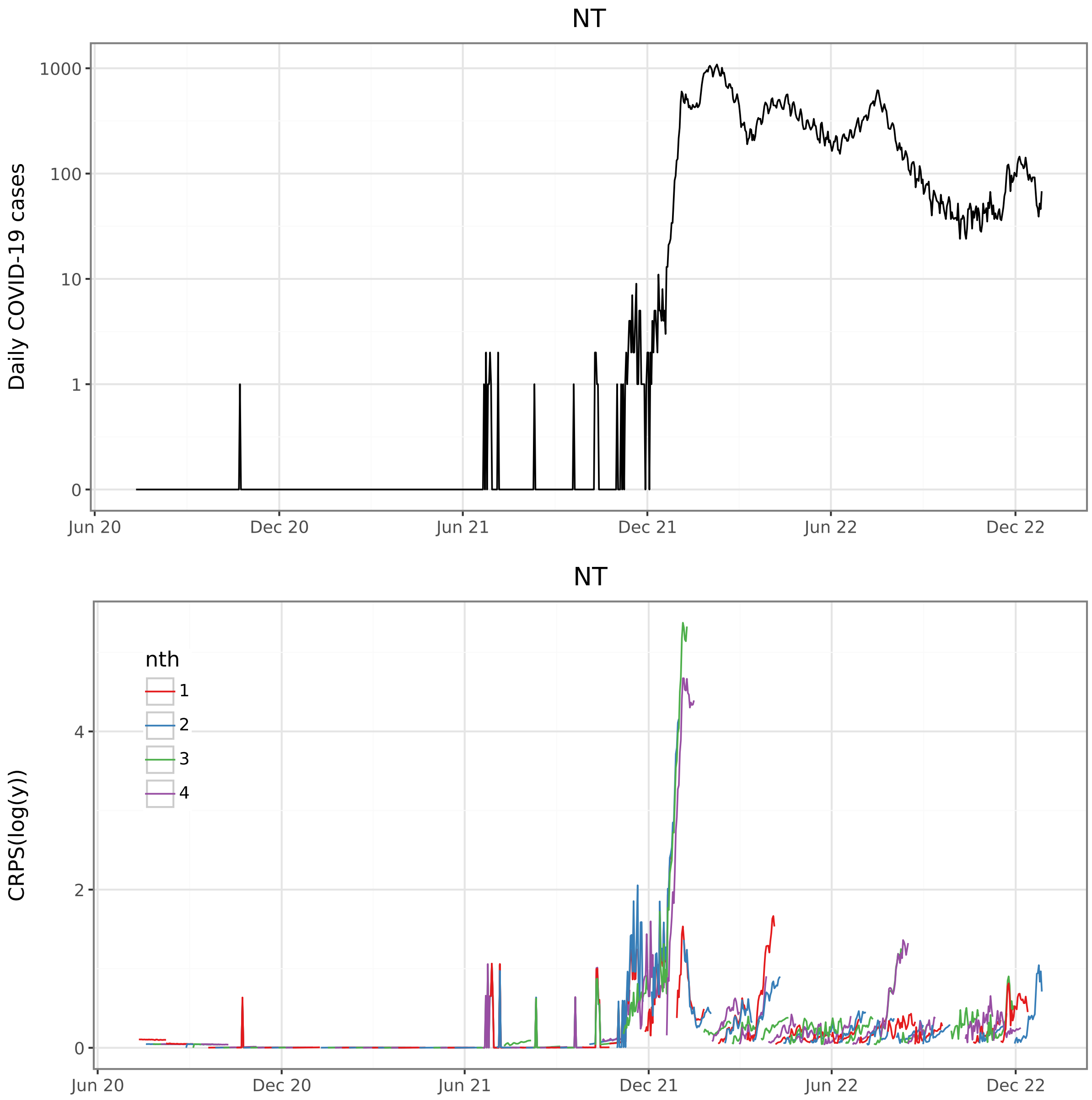


Figure S23: NT daily cases and CRPS.

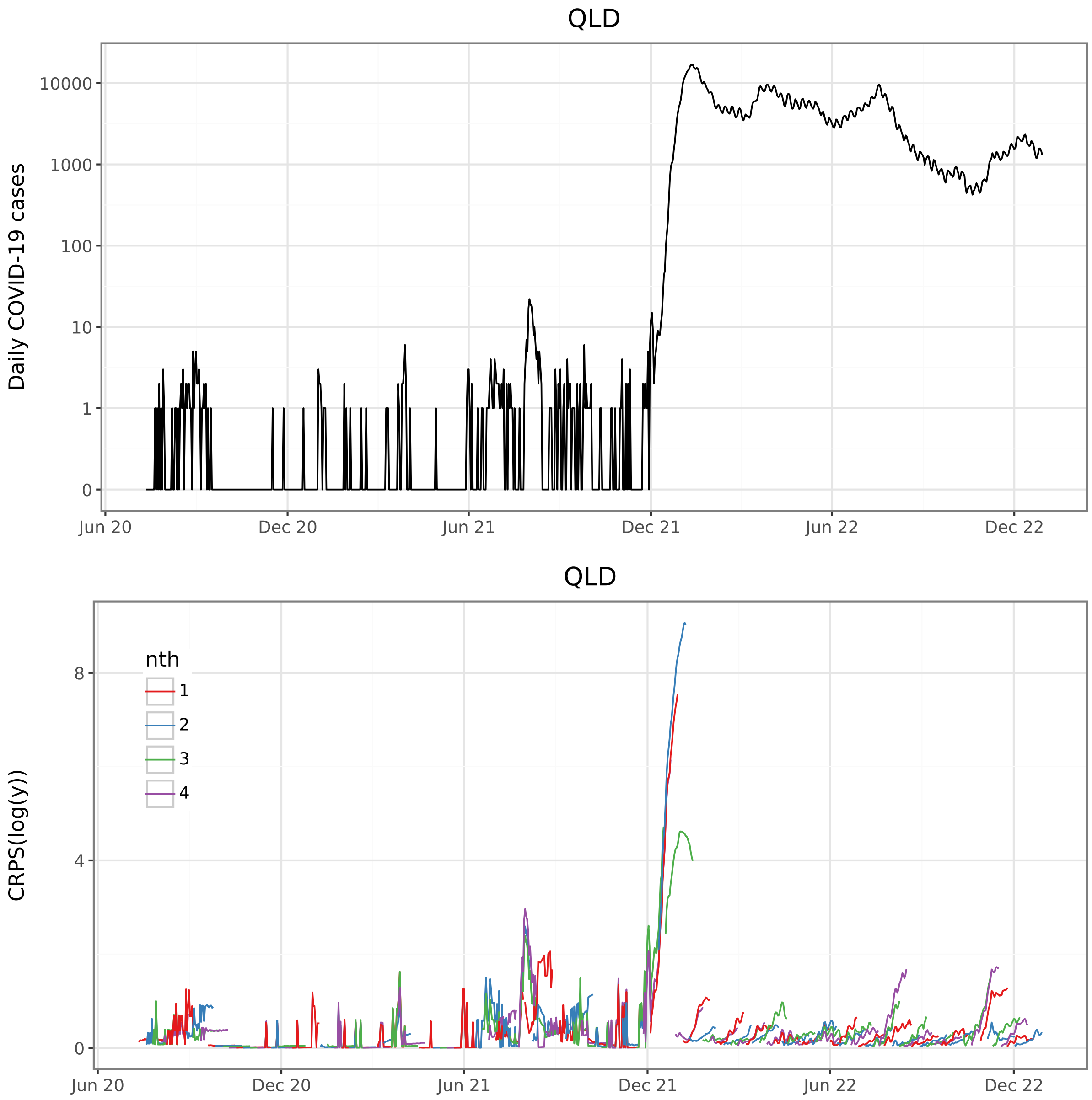


Figure S24: QLD daily cases and CRPS.

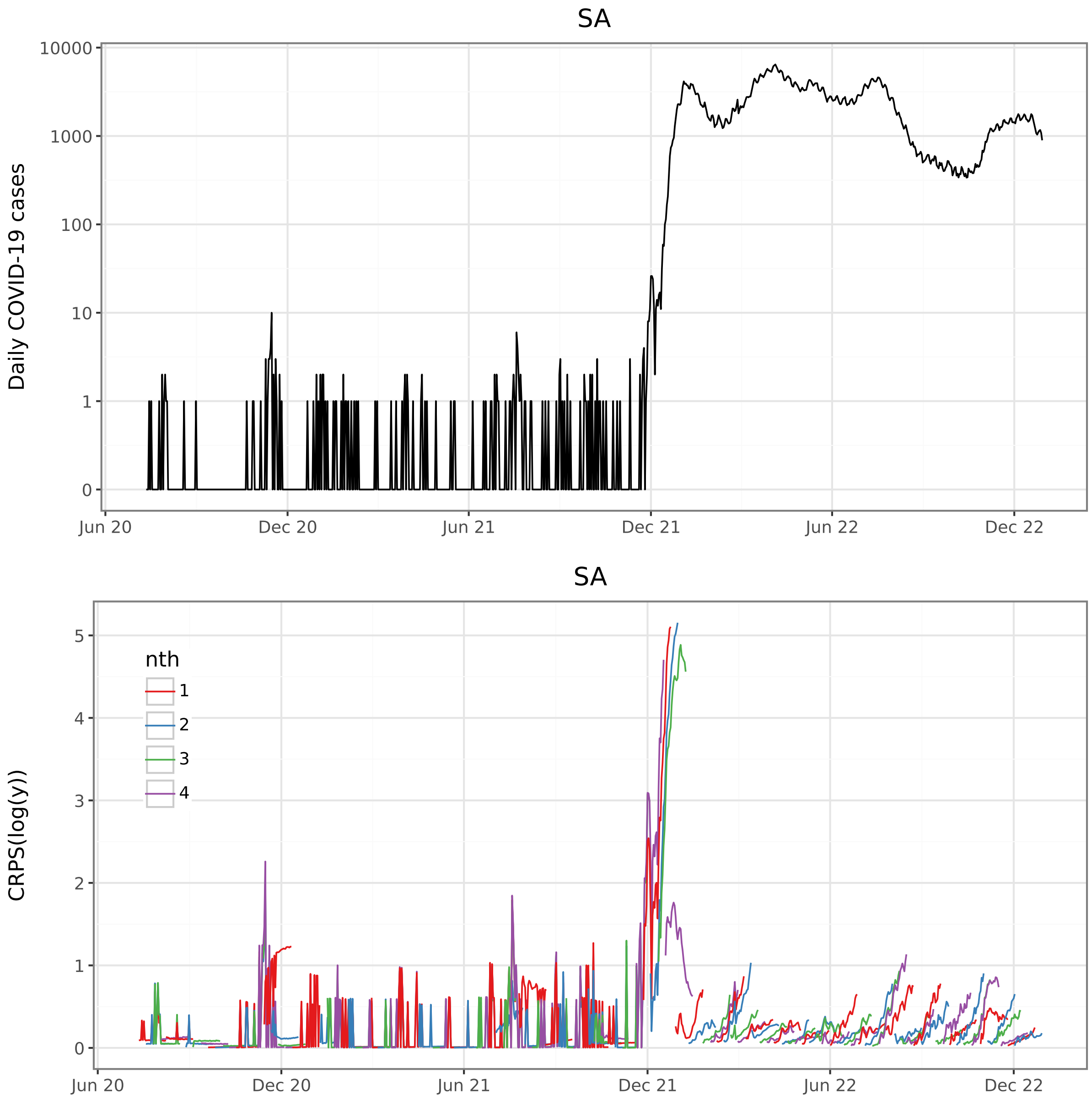


Figure S25: SA daily cases and CRPS.

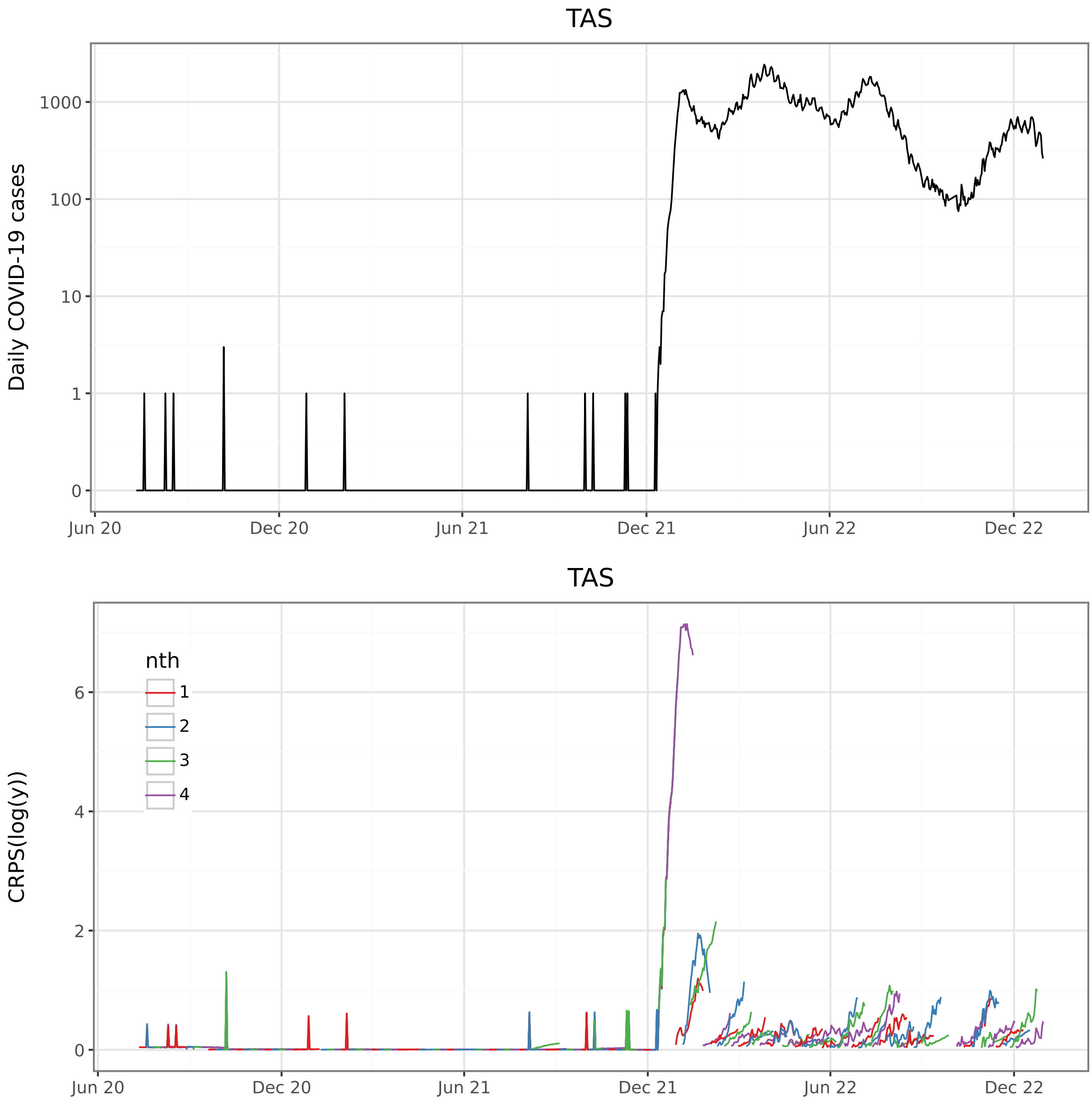


Figure S26: TAS daily cases and CRPS.

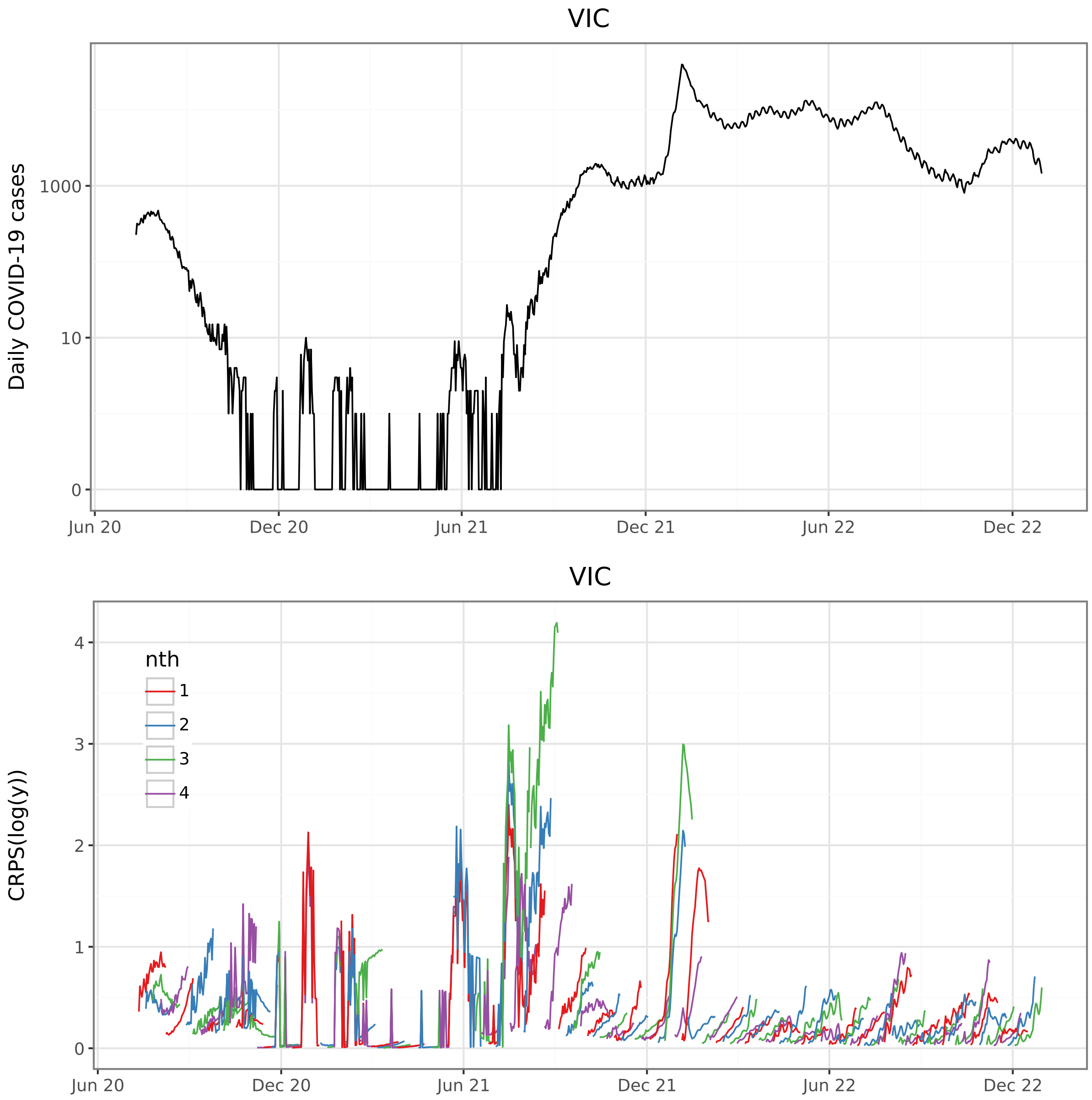


Figure S27: VIC daily cases and CRPS.

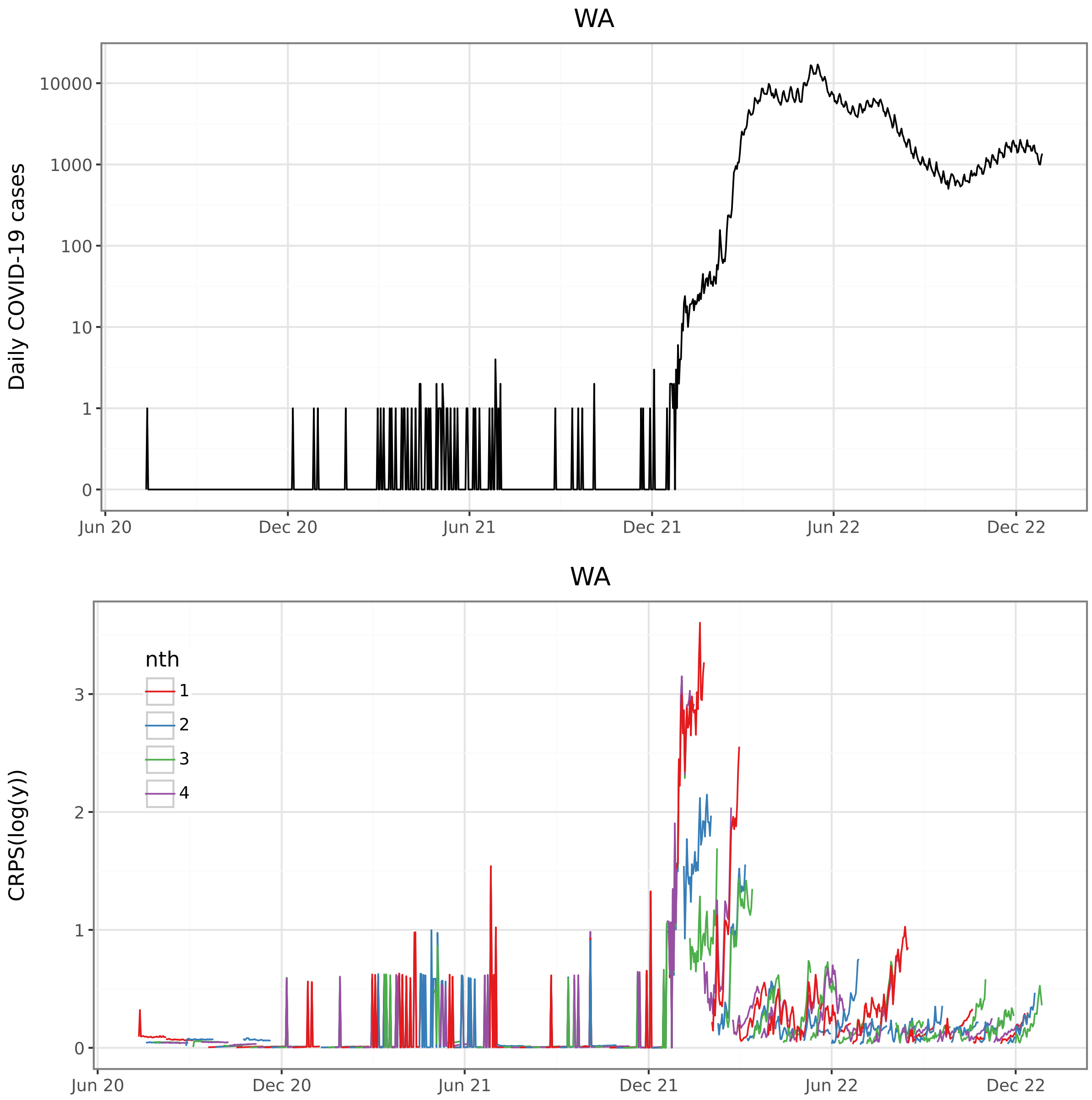


Figure S28: WA daily cases and CRPS.

#### S8 CRPS values for each dominant strain

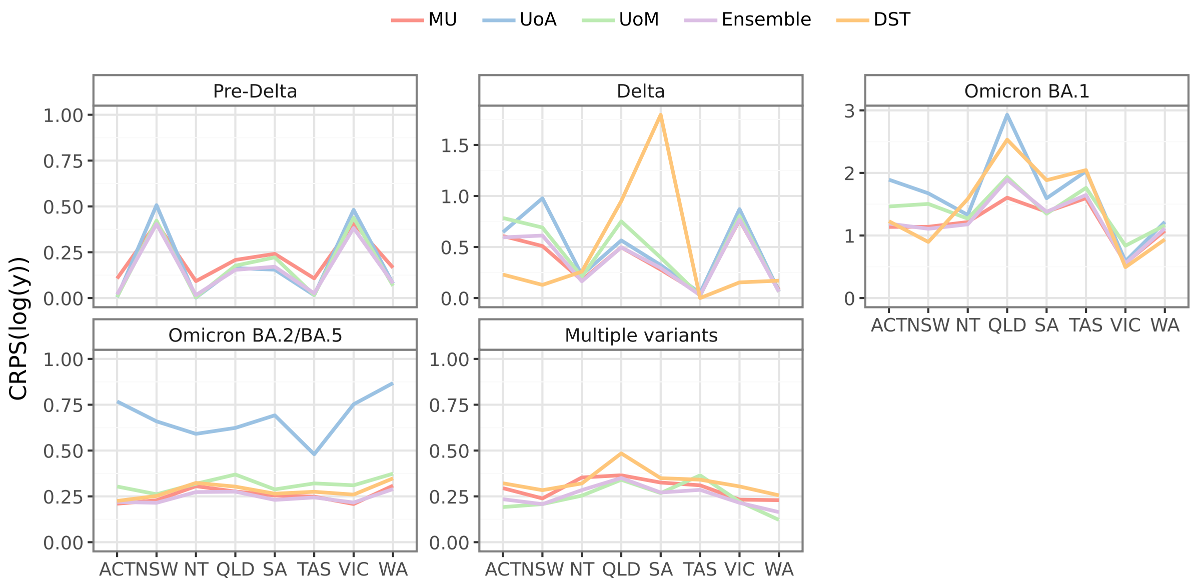


Figure S29: Mean CRPS for each model, and the ensemble, shown separately for each jurisdiction and for each dominant strain.
